# Effectiveness of active physiotherapy on physical activity level in community-dwelling stroke survivors: a systematic review and meta-analysis of randomized controlled trials

**DOI:** 10.1101/2023.04.21.23288899

**Authors:** Stéphanie Goncalves, Morgane Poitiers Le Bourvellec, Stéphane Mandigout, Noémie Duclos

## Abstract

**Background:** Stroke survivors are primarily physically inactive. Physiotherapy practice might represent professional support to improve this alarming lifestyle. However, evidence is scarce regarding the effectiveness of active physiotherapy on physical activity level in stroke survivors.

**Methods:** We conducted a systematic review and meta-analysis of randomized controlled trials (RCTs) according to the Preferred Reporting Items for Systematic Reviews and Meta-Analyses statement. Participants: Stroke survivors living in the community. Intervention: Any active physiotherapy, i.e., involving exercises that require voluntary effort by the patient. Outcome measure: Physical activity level.

**Results:** Out of 5590 identified references, 25 RCTs were eligible, and 21 had available data. Pooling resulted in a small significant effect size in favor of active physiotherapy (standardized mean difference (SMD) 0.22, 95% confidence interval (CI) 0.04 to 0.40, heterogeneity *I*^2^=65%), and medium significant effect when physical activity level was measured using an objective tool (SMD 0.48, 95% CI 0.03 to 0.92, *I*^2^=73%). In addition, meta-regression showed that 35% of the variance in trial outcome was explained by the measurement tool (objective or subjective) and 23% by age. Finally, none of the variances were associated with specific dosage in frequency, time, or exercise duration, nor with the severity of the disability.

**Conclusion:** Active physiotherapy seems effective in increasing objective physical activity among stroke survivors living in the community. However, the evidence supporting the efficacy of active physiotherapy was found to be of low certainty. Thus, further evidence is required.

**Registration PROPERO:** CRD42022315639.

## Introduction

Stroke is a major cause of long-term adult disability of growing importance. In the past three decades, stroke incidence increased by 70% ^1^. Simultaneously, improvements in acute care led to a mortality decline. As a result, the number of people living with stroke consequences (i.e., stroke survivors) in the community is rising ^2^. Among them, 4 out of 10 will have a recurrent stroke within 12 years after the first ^3^. Physical inactivity multiplies the risk of recurrence by five, making it one of the main risk factors ^4^. Unfortunately, community-dwelling stroke survivors often tend to be inactive ^5^. Stroke-related impairments frequently lead to a vicious circle of physical deconditioning and insufficient activity. For instance, they only walk half as many daily steps as healthy individuals ^6^. Thus, managing this population’s physical activity level (PAL) is urgently required.

Physiotherapists’ contribution to promoting physical activity is becoming increasingly obvious ^7^. They are associated with movement in the patient’s mind and appear well-placed to encourage physical activity ^8^. Their expertise enables them to tackle perceived barriers to physical activity and strengthen key facilitators such as walking ability, mobility, balance, or endurance, which are positively associated with long-term adherence to physical activity ^9^. Moreover, while physiotherapists play an essential and widely recognized role in the early stages of stroke recovery, evidence that their intervention is relevant in the chronic phase is growing ^10^.

In recent years, six systematic reviews have examined the effectiveness of interventions to improve PAL in individuals with stroke ^11–16^. However, most authors emphasized the impossibility of concluding due to the heterogeneity of interventions, which could encompass general lifestyle advice and exercise programs ^11–15^. Therefore, they recommended that research independently investigate each component of the contributions of the complex interventions ^11^. Previous research suggested that the PAL of various populations undergoing outpatient physiotherapy (e.g., people with Parkinson’s disease, low back pain, or chronic obstructive pulmonary disease) increases when physiotherapists focus on active treatment modalities ^17^. According to the most recent international stroke guidelines, the active aspect of physiotherapy treatment should be emphasized ^18^.

Two modalities in physiotherapy treatments can be distinguished: passive physiotherapy and active physiotherapy. Passive physiotherapy implies no form of physical activity defined as any skeletal muscle contraction resulting in energy expenditure ^19^, such as education or stretching. Conversely, active physiotherapy is defined as any “form of exercise carried out by the patient, under direct supervision from the physiotherapist or not, and requiring a voluntary effort on the part of the patient” (e.g., functional task training, circuit class therapy, aquatic therapy, or gait training) ^20^. With this in mind, one systematic review has examined active interventions solely to increase stroke survivors’ PAL ^16^. The authors focused on exercise programs that complied with moderate to vigorous physical activity guidelines. However, they found insufficient evidence of their beneficial effects on PAL ^16^, suggesting that other types of active treatment should be investigated.

Hence, the proposed research questions of this systematic review with meta-analysis were:

1. Is active physiotherapy effective in increasing the PAL of stroke survivors living in the community?
2. What are the effects of active physiotherapy interventions across their characteristics (e.g., type, duration, intensity), population profiles, and physical activity measurement tools?

## Method

The automatic tool “Cochrane Handbook for Systematic Reviews of interventions” (Version 6.2,2021) and the Preferred Reporting Items for Systematic Reviews and Meta-Analyses (PRISMA) updated guidelines were used for this work (Supplemental Material) ^21^.

This systematic review was registered prospectively on PROSPERO (ID: CRD42022315639).

### Electronic searches

Five core electronic databases were reviewed from inception to March 16, 2022: Medline via PubMed, Physiotherapy Evidence Database (PEDro), Excerpta Medica Database (EMBASE), Cumulative Index to Nursing and Allied Health Literature (CINHAL) via EBSCOhost, and Central (via the Cochrane Library). No language or date restrictions were applied.

Initially, the search strategy was developed for Medline (Supplemental Material). The concepts chosen referred to the acronym PICOS. In addition, we looked for synonyms in previous peer-reviewed publications and Cochrane reviews on a related topic and the National Library of Medicine Medical Subject Headings. Keywords and synonyms were combined with Boolean operators. Finally, we tested the strategy using a gold standard set developed during preliminary searches and adjusted it with an iterative process and the assistance of a librarian. Next, this search strategy was adapted to the syntax and subject headings of the other databases (Supplemental Material). We searched unpublished literature such as conference papers, abstracts, or presentations. Finally, we consulted clinicaltrials.gov and PROSPERO for ongoing or recently completed trials and systematic reviews.

### Eligibility criteria

#### Types of study

All the Randomized Controlled Trials (RCT), Quasi-RCT, and Controlled Clinical Trials that investigated the effectiveness of active physical therapy were included.

#### Types of participants

Studies included adult patients after the first or recurrent stroke, currently living in the community. Studies with participants who had a transient ischemic attack were excluded.

#### Types of interventions

Interventions involving exercises that require a volitional effort by the patient were included ^20^. We considered not only face-to-face and group approaches, but also remote interventions. In the case of remote intervention, we distinguished between interventions consisting of general advice on physical activity (studies not included) and a specific unsupervised exercise program (studies included).

When the physiotherapist was not mentioned but the specified program could be prescribed or implemented by a physiotherapist, the study was included. We included technological interventions but not pharmacological and neuromodulation interventions.

Interventions that do not require the patient’s physical effort (e.g., education, stretching) were excluded.

#### Types of comparators

Passive treatment (e.g., massage, education, manual therapy) or less active than the intervention, waitlist, no intervention, or usual care if we have enough knowledge of active components of this comparator, were included. When the difference between interventions was not the active ingredient, the trial was excluded.

#### Types of contexts

This review included therapeutic settings such as outpatient and community settings (private practice, center-based, or home-based). When the first intervention session occurred in an inpatient setting, while the remainder occurred in an outpatient setting, the study was included.

#### Types of outcomes

The primary outcome of this review was the PAL. Its measurement could be objective (i.e., with an activity monitor, accelerometer, or pedometer) or subjective (i.e., self-reported questionnaire, diaries), whatever the duration (after the intervention or onward).

Contrariwise, studies reporting solely on walking capacity or ability to perform activities of daily living were excluded.

### Selection of eligible articles and data extraction

The online platform COVIDENCE (Veritas Health Innovation, Melbourne, VIC, Australia), a relevant tool for rigorous systematic reviews ^22^, was used throughout the process. The results were imported into the ZOTERO bibliographic management software to detect and remove duplicates. The studies were selected with COVIDENCE, using predetermined eligibility criteria, appearing on a predesigned form. The screening was conducted by two reviewers independently of each other (SG, ML or ND) by reading the title and summary first, followed by the complete text, to determine the final inclusion.

Disagreements were discussed with a third examiner (SG, ML or ND) to reach a consensus if necessary.

Two reviewers (SG and ML) independently extracted data, and any disagreement was discussed until achieving a consensus. The data extraction form was based on the Cochrane Collaboration’s Checklist ^23^ and was tested with a few studies beforehand.

Study details and sample attributes were reported. Relying on a recent Cochrane meta-analysis, active physiotherapy interventions were categorized into functional task training, musculoskeletal, cardiopulmonary, and multi-component intervention, when at least two were mixed ^24^. In addition, the intervention was described using the FITT principles (Frequency, intensity, time, and type). Next, the outcomes were reviewed at the endpoint and follow-up. When studies used multiple outcomes, the most objective was chosen, and in the case of several objective measures, the most common across studies was retrieved.

When studies reported multiple follow-ups, the most recent follow-up after the intervention was extracted to capture its immediate effects.

For studies with missing information, we contacted the corresponding author.

### Risk of bias assessment

Two reviewers (SG and ML) judged independently, with COVIDENCE, the risk of bias (RoB) of each included study with the RoB tool for RCTs (version 1) from the Cochrane Collaboration^25^. To derive an overall summary RoB judgment for each trial (at low, unclear, or high RoB), we used the Cochrane suggested framework ^25^. As double blinding should not be considered a criterion for evaluating bias risk in physiotherapy research ^26^, item 3 was not considered by the overall judgment. Consequently, six criteria out of seven rated as low risk of bias defined a study as having low RoB. A discussion between the two authors (SG and ML) resolved disagreements, and the third reviewer (ND) mediated if necessary.

### Certainty of evidence

The overall certainty of evidence i.e., confidence in the effect estimates, was assessed based on the Grading of Recommendations Assessment, Development, and Evaluation (GRADE) approach^27^.

### Statistical analysis

Strict inclusion criteria allow consideration of trials that are sufficiently clinically homogeneous to enable data pooling. Accordingly, meta-analysis was performed using the ‘meta’ and ‘metafor’ packages of R (R Foundation for Statistical Computing, Vienna, Austria; available at http://www.R-project.org/; version 4.2.1) and the support of a statistician experienced in meta-analysis.

We used Wan’s Method to deal with missing standard deviation or mean values and convert reported statistics to required ones ^28,29^.

The effect of active physiotherapy on PAL was measured through a random-effects inverse variance meta-analysis to pool weighted standard mean differences (SMD) with their 95% confidence intervals (95%CI). The restricted maximum-likelihood method (REML) was used for all analyses to estimate between-study variance. When studies had more than one relevant arm, we included group interventions as separate comparisons within the meta-analysis.

Findings were presented using forest plots and tables. Effect sizes (i.e., SMD) were interpreted following Cohen’s magnitude criteria for rehabilitation treatment effects: d = 0.14–0.31 “small” effect size; d = 0.31–0.61 “medium” effect size; and d > 0.61 “large” effect size ^30^.

We quantified heterogeneity with I² statistic ^31^, for which values between 0% to 40%, 30% to 60%, 50% to 90%, and 75% to 100% were considered respectively to imply no important, moderate, substantial and considerable statistical heterogeneity ^32^.

Subgroup and meta-regression analyses were performed to identify differences in active physiotherapy on PAL and sources of heterogeneity. Heterogeneity across included studies was assessed using the χ² test for heterogeneity with a 5% level of statistical significance ^33^. For each moderator variable on population (age, sex, time since stroke, clinical phase of stroke, impairment severity, balance classification, gait classification), intervention (type, supervision, frequency, duration, intensity), and outcome measurement (physical activity level measurement tool), the meta-regression analysis helped to quantify the amount of heterogeneity accounted for (R²). Subgroup analysis involves splitting data. Therefore, regarding age, we used 65 years, which is a commonly used cut-off point to distinguish younger from older persons. Concerning time since stroke, we arbitrarily applied a 3-year cut-off. Further, regarding the intervention, frequency of sessions per week was divided into ≤3 and >3, and duration of the intervention was split into <12 weeks, between 12 and 24 weeks, and > 24 weeks. In addition, we used literature-based thresholds for other classifications ^34,35^.

We investigated publication bias using a funnel plot and assessed its asymmetry with the Pustejovsky test ^36^. A p-value < 0.05 was considered statistically significant.

Two sensitivity analyses were performed to assess the robustness of our main findings. First, we performed a leave-one-out analysis, repeating the main analysis and excluding one study each time. Then, we analyzed only the study with an overall low RoB, looking at methodological quality.

## Results

### Study selection

A total of 5590 references were drawn from the electronic search strategy after removing duplicates. Among 82 full-text eligible articles, 25 were included in the qualitative synthesis. Among these 25 trials, 21 had data that could be quantitatively analyzed. Supplemental Material contains the list of excluded studies and reasons for exclusion. A PRISMA flow diagram illustrates each step in the review process (Figure 1). We contacted the authors of 5 trials to obtain more information, and only one answered.

**Figure 1:**
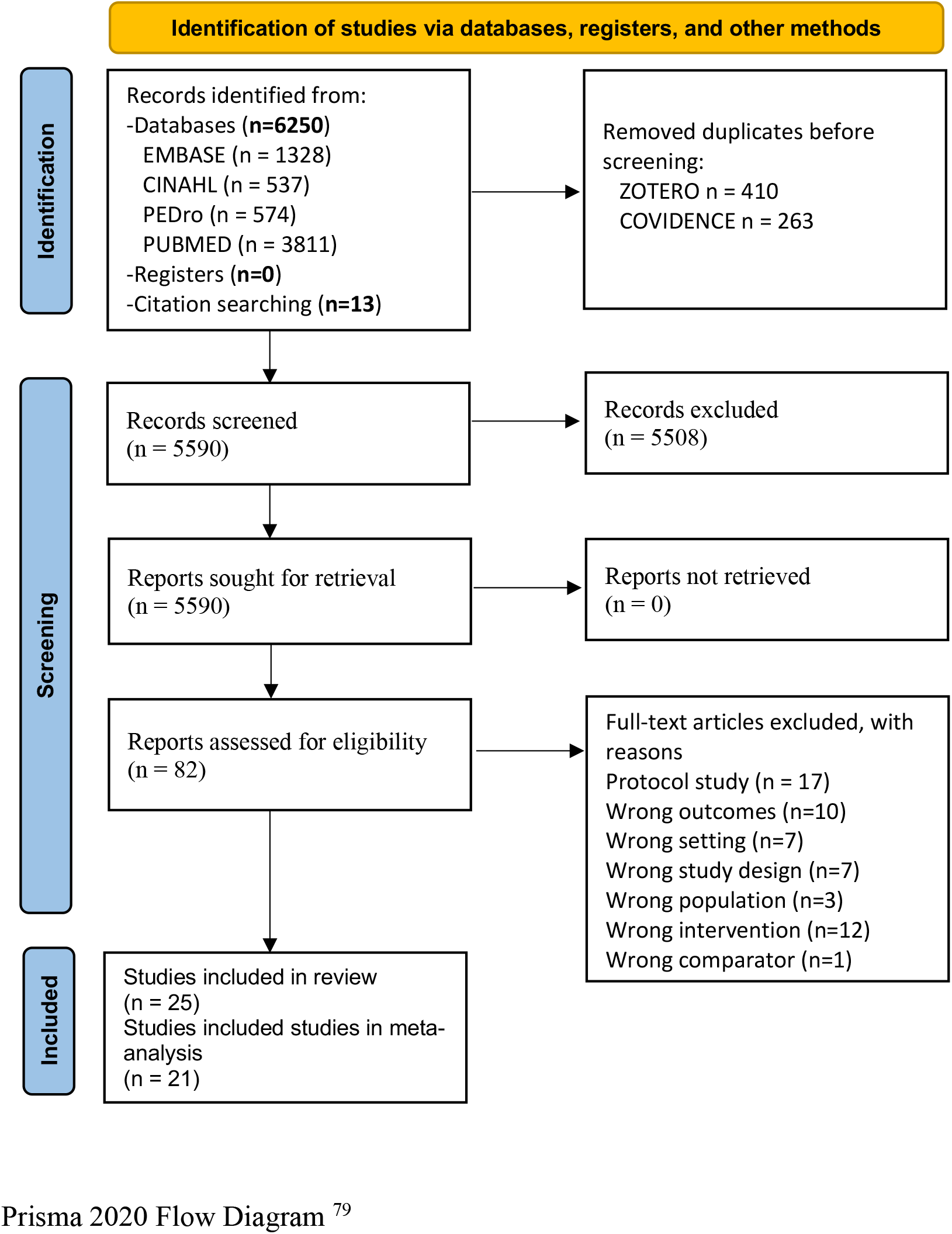
Flow Diagram.

### Study characteristics

The overall characteristics of the included trials are summarized in Table S1 in Supplemental Material. Ten studies were carried out in Europe ^37–46^, eight in America ^15,46–52^, four in Asia ^53–56^, and two in Oceania ^57–59^.

#### Design

Among the 25 trials, 23 had a parallel group design, and two had a cross-over design. However, data from the cross-over trials were unavailable at the end of the first period, so they could not be integrated into the meta-analysis.

#### Participants

The 25 studies involved 2448 stroke survivors, including 948 (39%) women. The sample size varied from 20 to 380, with a median of 56 participants. The mean age of participants ranged from 52 to 77.7 years, with a median of 60 years. Regarding time since stroke, six studies included participants in the subacute phase^37,38,42,53,57,60^ and 19 study participants in the chronic phase (range from 0.7 to 8.5 years). The severity of the participant’s motor impairment was reported by 12 studies: nine included participants with a mild deficit ^37,39–42,50,50,57,59^, two with a moderate deficit ^46,56^, and one with no deficit ^38^. The cognitive function was assessed in eight studies: six reported no deficit ^37,39,41,51,55,58^, and two had a mild deficit ^42,56^. Moreover, comfortable walking speed was evaluated in 15 studies with an average ranging from 0.38 to 1.1 m/s. Based on the Perry et al. categorization ^34^, participants could be characterized as limited community ambulators in seven studies ^44,47,49,54,57–59^, full community ambulators in six ^37,40,45,48,50,61^, and home ambulators in one ^46^. Balance was evaluated in eight trials ^37,39–41,46,49,54,58^, and participants were considered as independent for main transfers in five studies ^37,40,41,49,54^ and as requiring assistance in three ^39,46,58^.

#### Interventions characteristics

Six studies used cardiopulmonary interventions ^43,44,47,49,52,61^, one used musculoskeletal interventions ^54^, four used functional task training ^46,50,55,56^, and 14 studies used multi-component interventions ^37–42,45,48,51,53,57–60^.

The interventions were supervised, except in four studies that evaluated unsupervised programs set by a physiotherapist ^37,38,46,57^. Providers were physiotherapists ^40,42–45,50–53,55,56,58,59,61^, physiotherapy students ^59^, occupational therapists ^42,51,60^, trainers ^39,47,54^, exercise instructors ^51^, volunteers and qualified exercise instructors supported by a physiotherapist ^41^ and recreation therapists, educators, exercise therapists or other personnel with experience in healthcare or with stroke ^48^. Active physiotherapy was delivered at home in four studies ^37,38,46,57^, in a laboratory in five studies ^47,50,53–55^, a hospital or clinic in four ^40,42,43,59^, and in a community setting for the others ^39,41,48,51,52,58^. More than half of the interventions were group-based ^39–42,45,48,50,51,54,55,58–61^, with five involving circuit training ^41,45,50,58,59^.

Some interventions were based on existing programs, such as the Otago exercise program ^57^, WEEB (Weight-bearing Exercise for Better Balance) ^58^, HIFE (High-Intensity Functional Exercise program) ^42,45^ or FAME (Fitness And Mobility Exercise) ^48,51^. Equipment used for performing aerobic training in cardiopulmonary interventions was a cycle ergometer using either the lower limb or the upper limb ^43,44^, or a treadmill ^47,49,52,61^, one of which had a body-weight support ^49^. Among other technological devices, one study employed a Whole-Body Vibration platform (Gymna Fitvibe Medical System, Gymna Uniphy Pasweg, Bilzen, Belgium) ^54^ and another an over-ground robotic-assisted gait training device (Alter-G, Bionic Leg orthosis, Fremont, CA, USA)^46^.

The frequency of active physiotherapy sessions varied from one to seven times a week, on average, three times a week. Duration varied from 12 to 120 minutes, averaging 48 minutes per session. The intervention lasted from 2 weeks to 2 years with a median of 12 weeks. The intensity of the interventions was considered light in six studies ^38,54–57,60^, moderate in eleven ^39–41,43,45,48,50,51,53,58,59^, and vigorous in nine ^37,42,44,46,47,49,52,54,61^.

#### Control group characteristics

Two thirds of the control groups had passive or less active interventions. Two groups were not offered treatment (waitlist) ^48,49^, and the rest received usual care ^37–41,43,57,60^.

#### Outcome measures

Ten RCTs used an objective tool to measure PAL with an accelerometer ^39,46,50,53,56,61^, or a pedometer ^47,49,58,59^. Fifteen RCTs used a subjective measure such as the Frenchay Activity Index (FAI) ^41–43,54,55^, International Physical Activity Questionnaire ^37,60^, Human Activity Profile-Adjusted Activity Scores ^57^, Physical Activity Scale for Individuals with Physical Disabilities ^51^, Physical Activity Scale for the Elderly ^38,45^, Physical Activity Scale ^44^, Community Healthy Activities Model Program for Seniors ^48^, Yale Physical Activity Survey ^52^, and outdoors walking time per day report ^40^. In addition, PAL was assessed as a secondary outcome in half of the studies, after physical or functional outcomes such as walking ability ^37,42–47,53–55,57,58^.

### Meta-analysis

A meta-analysis of 21 trials was performed, including 1834 participants. Two trials had three arms with two active intervention groups ^54,55^. We included each pair-wise comparison in the meta-analysis.

Overall, pooling resulted in a small significant effect size in favor of active physiotherapy (standardized mean difference (SMD) 0.22, 95% CI 0.04 to 0.40, heterogeneity *I*^2^=65%) compared to the control group (Figure 2). With a *Z* value of 2.41 and a *P* value of .02, active physiotherapy is, on average, effective in increasing PA level in stroke survivors.

**Figure 2:**
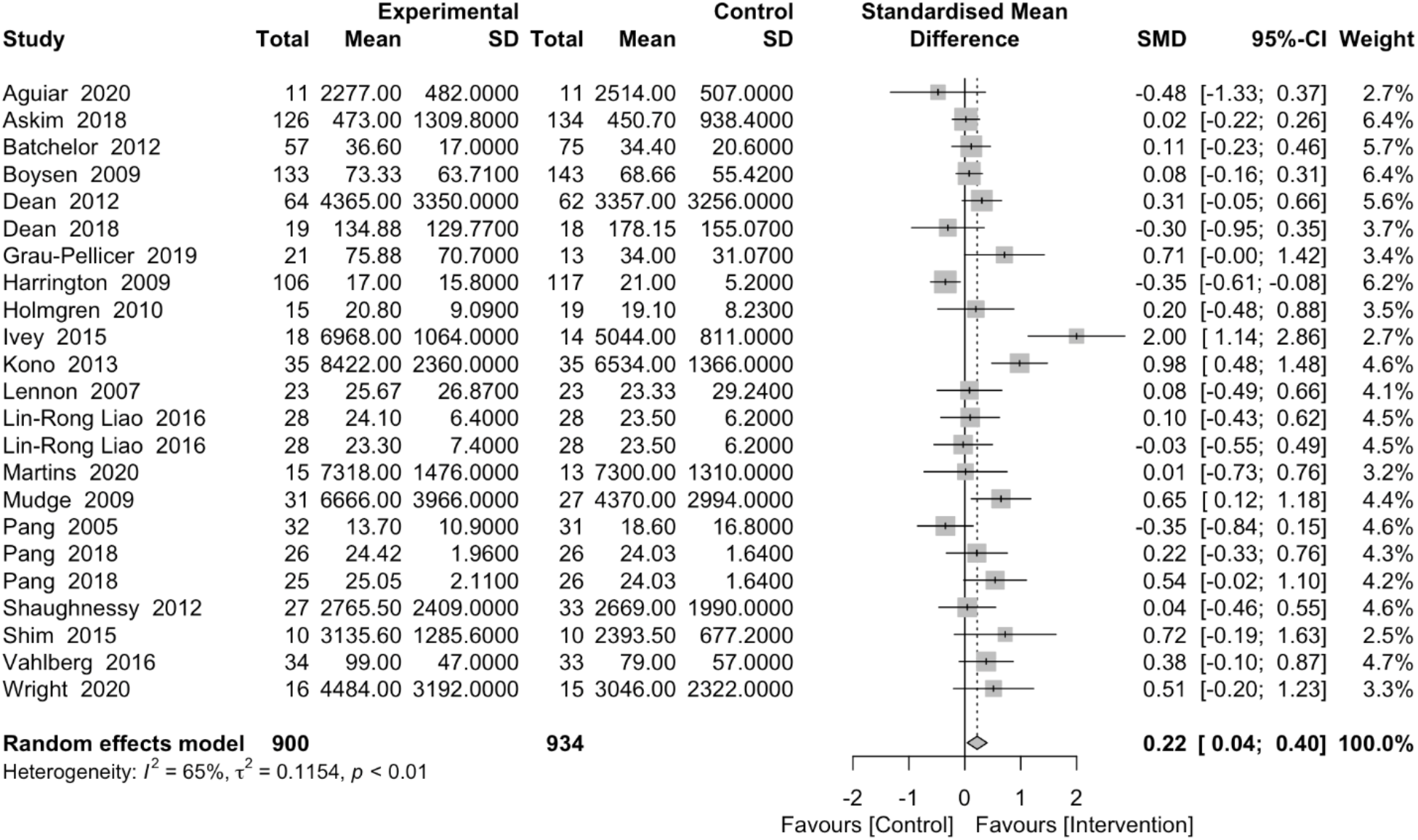
Overall meta-analysis (post-intervention)

A categorical pre-specified subgroup analysis was performed and is summarized in Table S2 in Supplemental Material.

When PAL was measured with an objective measure, active physiotherapy had a significant medium effect (9 studies, 424 participants, SMD 0.48, 95% CI 0.03 to 0.92, *I*^2^=73%). In contrast, with subjective measures the effect was small and non-significant (14 studies, 1410 participants, SMD 0.06, 95% CI -0.08 to 0.21, *I*^2^= 34.9%).

Active physiotherapy had a significant medium beneficial post-intervention effect when compared to less active or passive intervention (16 studies, 826 participants, SMD 0.34, 95% CI 0.10 to 0.57, heterogeneity *I*^2^=61.8%), and a non-significant effect when compared with usual care (7 studies, 1008 participants, SMD -0.01, 95% CI -0.19 to 0.19, heterogeneity *I*^2^=50.4%).

A significant medium SMD in favor of active physiotherapy was found for the subgroup of studies that included stroke survivors under 65 years (13 studies, 558 participants, SMD 0.39, 95% CI 0.10 to 0.68, heterogeneity, *I*^2^=62.8%), without subgroup difference between younger and older participants (*P*=.06). When the participants had a mean time since stroke under three years, active physiotherapy had a medium significant effect on PAL (7 studies, 578 participants, SMD 0.23, 95% CI 0.02 to 0.44, heterogeneity, *I*^2^=11.8 %), whereas when participants had a mean time since stroke over three years, active physiotherapy had a non-significant effect on PAL (12 studies, 627 participants, SMD 0.21, 95% CI -0.09 to 0.51, heterogeneity, *I*^2^=65.5%)

Among subgroups of types of interventions, functional task training had a significant medium beneficial effect on physical activity level, with no heterogeneity (5 studies, 182 participants SMD 0.38, 95% CI 0.08 to 0.67, *I*^2^=0%), whereas other interventions had a non-significant effect on PAL (Table S2 in Supplemental Material).

Results from the meta-regression models identified the PAL measurement tool (*R*²=35.29, *P*=.03) and the age of participants (*R*²=22.5, *P*=.05) as significant moderators (Table S3 in Supplemental Material).

Meta-regression did not reveal a statistically significant relationship between SMD and the treatment dose (i.e., frequency, duration, and intensity) and between SMD and clinical characteristics such as impairment severity, balance, and gait classification (Table S3 in Supplemental Material).

Ten studies assessed the long-term effects of the intervention (576 participants). The meta-analysis showed a non-significant effect (SMD 0.06, 95% CI -0.21 to 0.33, I^2^= 54.2%) of the experimental intervention compared to the control (Figure S1 in Supplemental Material).

### Risk of Bias

A summary of the RoB assessment is provided in Figure S2 in Supplemental Material. RoB was low in 76% of studies for allocation concealment, 81% for masking of outcome assessors, 71% for incomplete outcome data, 52% for selective outcome reporting, and 81% for other sources of bias. Overall, nine studies were at low RoB ^37,38,42,55,57–59,61^, three at unclear RoB ^43,51,53^, and ten at high RoB ^39–41,45–47,50,52,54,56^.

### Sensitivity analysis

The first sensitivity analysis (Table S2 in Supplemental Material), which restricts meta-analysis to studies at lower RoB, found a statistically significant result (9 studies, 1011 participants, SMD 0.15, 95% CI 0.03 to 0.27, I^2^=17.4%). The second sensitivity analysis exploring the impact of excluding each trial on the results is summarized in Figure 3.

**Figure 3:**
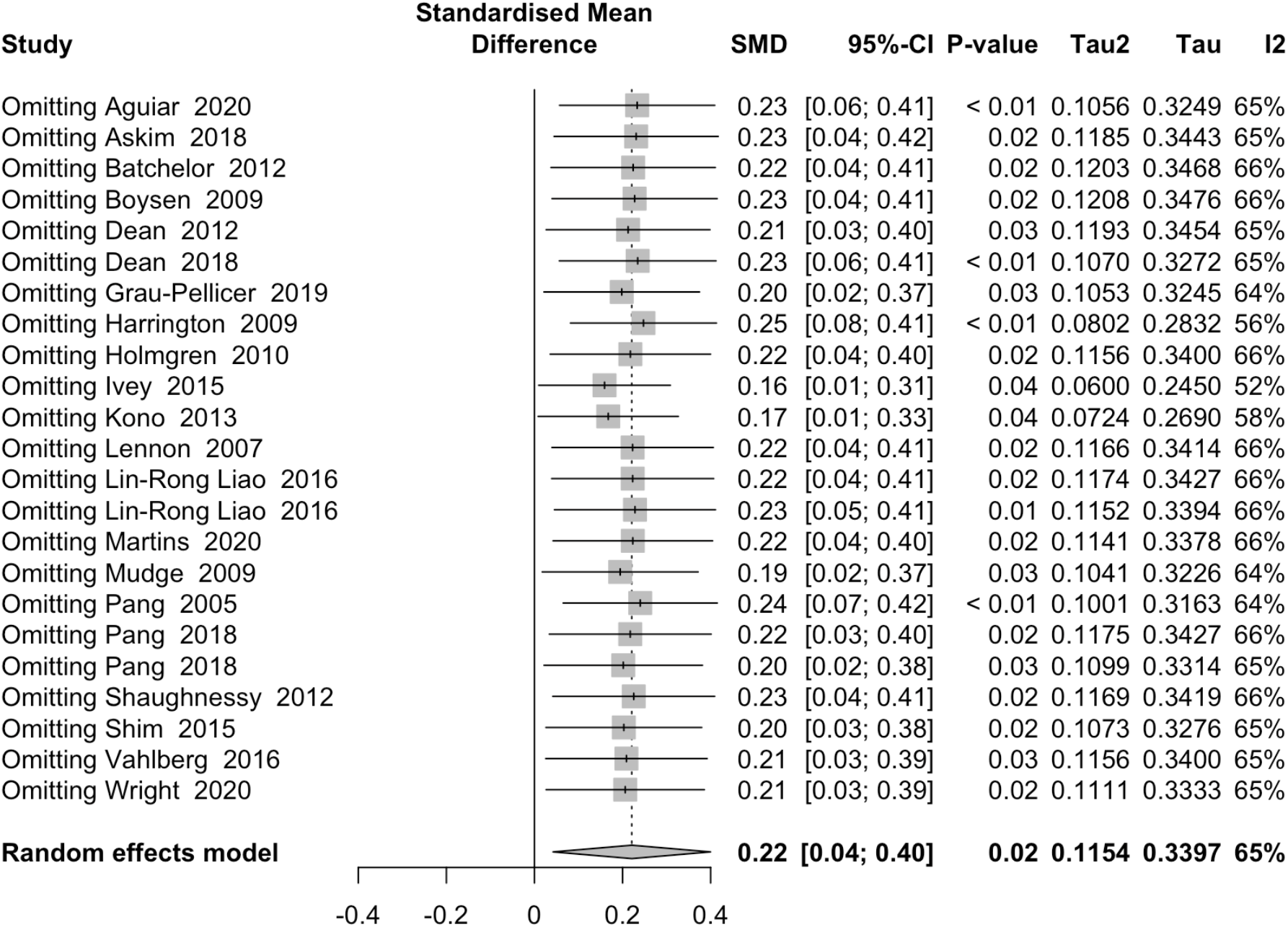
Leave one out analysis.

Our findings suggest a low risk of publication bias based on examination of the funnel plot and underpinned by the statistical method (p>0.05) (Figure 4).

**Figure 4:**
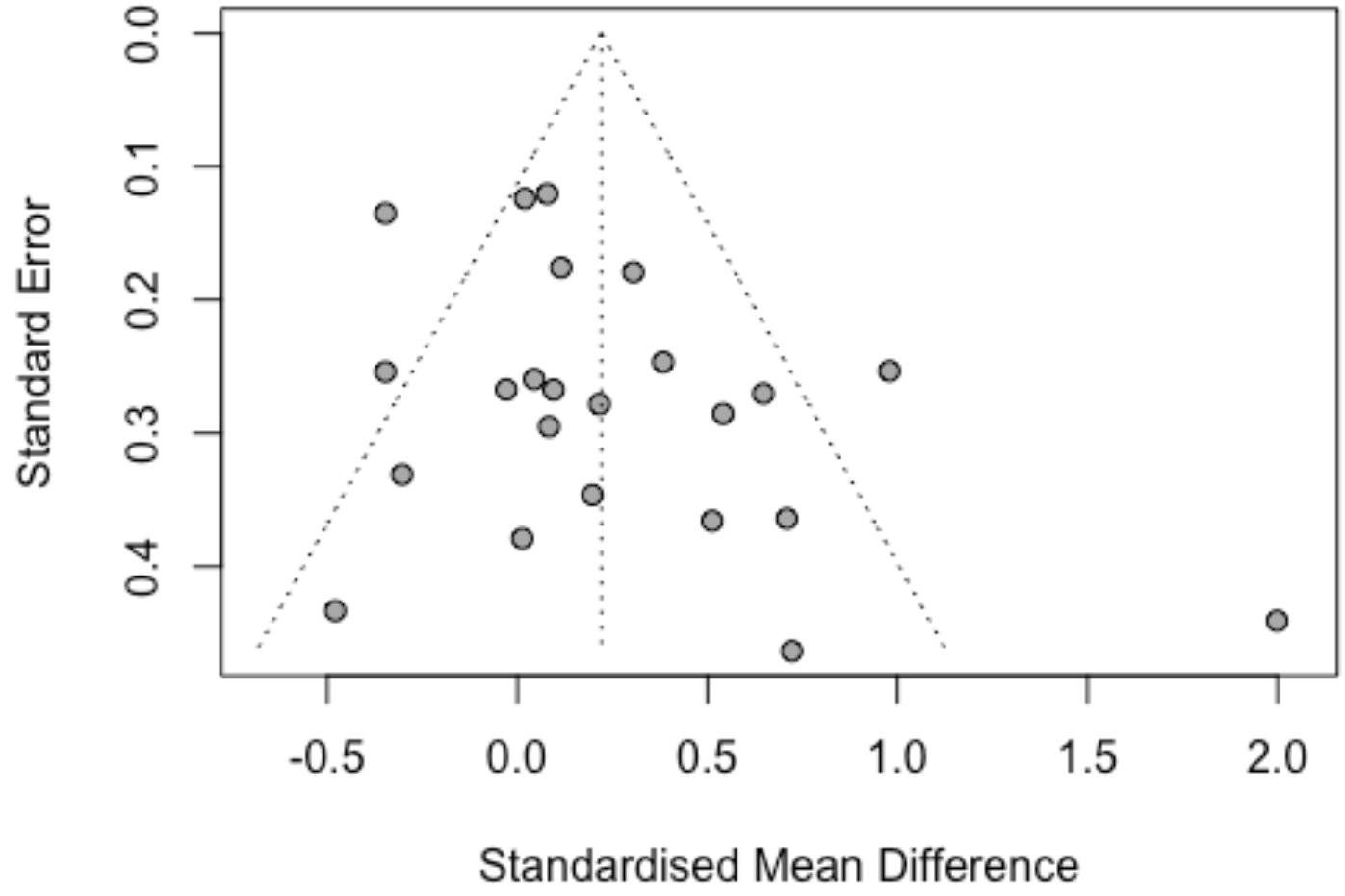
Funnel Plot. P value publication bias = 0.080

### Certainty of evidence

#### Study limitations

58% of the studies were at unclear to high RoB. However, our first sensitivity analysis, restricting meta-analysis to studies at lower RoB, found a similar confidence interval, indicating the robustness of our findings. Additionally, the second sensitivity analysis exploring the impact of excluding each trial on the results showed remaining consistency with the overall findings. As a result, we did not downgrade PAL for study limitations.

#### Imprecision

Upper and lower boundary of the confidence interval represents a beneficial effect of active physiotherapy on PAL. However, minimal clinically important difference (MCID) thresholds in stroke survivors were not determined in any study for PAL outcome measures. Therefore, PAL was downgraded by one level for imprecision.

#### Inconsistency

The I^2^ value of 65 % indicates substantial statistical heterogeneity. Subgroup analyses provided potential explanations but not complete ones. PAL was consequently downgraded due to inconsistency.

#### Indirectness

Evidence was downgraded due to indirectness because participants appear to have predominantly mild to moderate impairment and to be younger than stroke patients who could receive effects from these interventions. Therefore, our results were only partially applicable to our question and were difficult to generalize to older patients or those with severe impairments. Accordingly, we downgraded PAL for indirectness.

#### Publication bias

We found a low likelihood of publication bias with the funnel plot and statistical method (p>0.05). Therefore, the evidence was not downgraded for publication bias.

Finally, neither the dose-response gradient, nor the magnitude of effect nor confounding could upgrade the certainty of evidence.

In summary, according to the GRADE approach, the certainty of evidence was judged very low.

## Discussion

### Summary of main results

To the best of our knowledge, this is the first systematic review and meta-analysis of RCTs regarding the effectiveness of active physiotherapy on PAL in community-dwelling stroke survivors. Overall, our findings indicated a small positive and significant effect of this intervention compared with the control group immediately after the intervention. These results support the awareness of international organizations, including the World Health Organization and the World Physiotherapy (i.e., the international voice of physiotherapy), of the fact that physiotherapists play a pivotal role in PA promotion ^62–64^. In addition, health professionals are recognized as significantly impacting how active stroke survivors are ^65^. However, the beneficial effect was not maintained at follow-up, reflecting difficulties in promoting long-term sustainable physical activity.

Interestingly, our subgroup analysis found that the studies using objective methods demonstrate a medium effect compared to subjective ones. The meta-regression indicated that 35.29 % of the heterogeneity was accounted for by the PAL measurement tool, which therefore represents the strongest predictor of overall effect. Objective methods such as a pedometer or accelerometer are considered to provide more accurate measures of PAL than subjective ones. In contrast, self-report measures could suffer from recall and desirability bias on the part of the participants ^66^. This suggests a medium effect of active physiotherapy on stroke survivors’ PAL. In addition, among the subjective tools used in our review, PAL was assessed with the FAI in five studies. However, a recent systematic review on measurement properties of self-report PAL assessment tools for patients with stroke pointed out, in this questionnaire, the conflict between items on PAL and items on the performance of ADL ^67^. The assessed criterion of behavior as complex as a physical activity should be clarified. Nevertheless, it should be noted that the objective tools do not provide information about the circumstances, such as leisure, professional, and transport activities or domestic activities allowing precise and complete measurement of PAL. Furthermore, many recommendations are based on the relationship between self-reported physical activity and health ^68^. Therefore, subjective measures should still not be neglected. Finally, these results must be taken cautiously as few objective tools used in this review have been validated in this pathological population ^39,46,47,50,53,56,58,59,61^.

Next, this research found a medium post-intervention effect in improving PAL when active physiotherapy is compared to less active or passive intervention. Conversely, no effect was demonstrated when intervention was compared to usual care. This could be explained by the “black box of usual care” ^69^ and insufficient information about activity performed in the control group ^70^. For instance, in the LAST study ^37^, the control group received 45 minutes of physiotherapy per week for up to six months, which exceeds the norm in many nations where access to physiotherapy in the chronic phase post-stroke is limited ^71^.

Another point of interest is that, regarding intervention groups, only functional task training had beneficial moderate effect on PAL, emphasizing the importance of one of the guiding principles for effective neurorehabilitation ^72^. Multi-component intervention had a non-significant effect, corroborating previous findings which suggest that interventions are more effective when they contain fewer components ^11^. This could also be justified by the population for which the programs were developed, such as the Otago exercise program ^57^, WEEB ^58^, HIFE ^42,45^, or FAME ^48,51^ for older people and not specifically designed for post-stroke patients. Finally, most interventions were group-based, supporting evidence from previous work considering that the opportunity to interact with other stroke survivors is the most predominant incentive for physical activity after stroke ^73^.

Afterward, active physiotherapy seems more effective among younger (<65 years old) than older patients (>65 years old). However, age contributes to increase in the likelihood of being physically inactive one year following a stroke ^74^. In contrast, the association between older age and a lower probability of undergoing rehabilitation has been reported in the literature ^75^, highlighting the urgent need to target this specific population with personalized approaches.

Finally, in contrast with earlier work ^76^, the meta-regression revealed a lack of dose-response relationship. One possible explanation could be insufficient reporting of stroke survivors’ adherence to prescribed exercises, as was suggested in an earlier systematic review ^77^. Particularly, adherence to prescribed exercises when exercise is unsupervised appears concerning. For example, in the LAST study ^37^, despite prior coaching only 43 to 64% of the participants reached the weekly prescribed exercise duration ^37^. Furthermore, substantial heterogeneity in the analysis prevents us from drawing conclusions about this relationship.

### Overall completeness and applicability of evidence

First, our review research question addressed stroke survivors regardless of age (18 and older) or impairment severity. However, we only found evidence concerning patients with a median age of 60, from 55 to 75, and mild to moderate impairment. So, evidence may not directly apply to older stroke survivors or patients with severe impairment. Studies have been conducted in several countries worldwide, with substantial variations in physiotherapy and health care systems, which may limit the extent to which these results can be applied internationally.

### Certainty of evidence

The certainty of evidence was downgraded to very low certainty of evidence for imprecision, inconsistency, and indirectness, leading to a cautious conclusion on our results on the effectiveness of active physiotherapy in increasing PAL among stroke survivors.

### Agreements or disagreements with other studies or reviews

Our results corroborate the findings of Kunstler et al. ^17^, even though the studies included in their review recruited more broadly any patient with a non-communicable disease, and not specifically stroke survivors. Their meta-analysis of continuous outcomes showed a small significant effect of active physiotherapy post-intervention, but not in the long term ^17^. Moreover, their meta-analysis of dichotomous outcomes showed that active physiotherapy doubled the odds of patients achieving the minimum recommended PAL.

To date, there have been six systematic reviews on interventions to improve stroke survivors’ PAL, but only one conducted a meta-analysis that could permit comparison with our work ^16^. However, the exercise schedule included in the meta-analysis should be at least moderate in intensity, making it difficult to compare their findings with ours.

### Strengths and Limitations

The strength of this systematic review and meta-analysis was the robust methodological process and the narrow inclusion criteria allowing performance of a meta-analysis. However, this review process had several limitations. Firstly, decisions to judge a study at low RoB even though masking participants and personnel were at high RoB could have introduced bias in review findings. In addition, our classification of types of interventions could have modified the results^24^. For instance, we classified walking on a treadmill as cardiopulmonary training due to the moderate intensity of this intervention, but it could have been classified as functional task training. In the same way, we classified circuit class therapy as multi-component training even though it could have been classified as functional task training or cardiopulmonary training since this model of physiotherapy delivery is provided in an intensive manner and is focused on functional tasks ^78^. Lastly, while several study authors were contacted, few responses were received. Thus, insufficient data from these studies led to their exclusion from the meta-analysis, which may have produced different findings.

### Perspectives

This research may have important clinical implications. Beyond rehabilitation of function, active physiotherapy could influence the physical activity patterns of stroke survivors living in the community. In the face of the vital need to tackle physical inactivity and its deleterious consequences, physiotherapists could play a significant public health role.

From a research perspective, our review highlights the importance of using objective measures to assess the effectiveness of active physiotherapy and the requirement of validated tools. Another challenge for future research will be stratifying patients according to age to better tailor the interventions. In addition, our findings clarify the need to better define the interventions in the control group, particularly active ingredients. Lastly, a few trials have assessed the effectiveness of active physiotherapy on PAL as the primary outcome, reinforcing the relevance for future RCT to prioritize outcomes of utmost importance, such as PAL.

## Conclusion

Our findings support the effectiveness of active physiotherapy to increase physical activity level in community-dwelling stroke survivors, with larger effect when measured with an objective tool than with a subjective one. Due to the very low certainty of evidence, definitive conclusions seem to be premature. This emphasizes the need for further investigation.

## Data Availability

Data available on request from the authors

## Acknowledgments

We thank for Dr Jean Joël Bigna Rim for the support in statistics

## Sources of Funding

This research was supported by the funds of the French Society of Physiotherapy (Société Française de Physiothérapie, SFP)

## Disclosures

The authors have no conflicts of interest to declare

## Notes

### Competing Interest Statement

The authors have declared no competing interest.

