## Supplemental M.2 for "Effectiveness of active physiotherapy on physical activity level in community-dwelling stroke survivors: a systematic review and meta-analysis of randomized controlled trials"

### Search strategy on Medline

(("Cerebrovascular Disorders"[MeSH Terms:noexp] OR "Basal Ganglia Cerebrovascular Disease"[MeSH Terms] OR "Brain Ischemia"[MeSH Terms] OR "Carotid Artery Diseases"[MeSH Terms] OR "Intracranial Arterial Diseases"[MeSH Terms] OR "Intracranial Embolism and Thrombosis"[MeSH Terms] OR "Stroke"[MeSH Terms] OR "Brain Infarction"[MeSH Terms] OR "stroke, lacunar"[MeSH Terms] OR "vasospasm, intracranial"[MeSH Terms] OR "Vertebral Artery Dissection"[MeSH Terms] OR "Hemiplegia"[MeSH Terms:noexp] OR "Paresis"[MeSH Terms] OR ("Stroke"[Title/Abstract] OR "poststroke"[Title/Abstract] OR "post-stroke"[Title/Abstract] OR "cerebrovasc*"[Title/Abstract] OR "brain vasc*"[Title/Abstract] OR "cerebral vasc*"[Title/Abstract] OR "cva"[Title/Abstract] OR "apoplex*"[Title/Abstract] OR "SAH"[Title/Abstract]) OR (("brain*"[Title/Abstract] OR "cerebr*"[Title/Abstract] OR "cerebell*"[Title/Abstract] OR "intracran*"[Title/Abstract] OR "intracerebral"[Title/Abstract]) AND ("ischemi*"[Title/Abstract] OR "infarct*"[Title/Abstract] OR "thrombo*"[Title/Abstract] OR "emboli*"[Title/Abstract] OR "occlus*"[Title/Abstract])) OR (("brain*"[Title/Abstract] OR "cerebr*"[Title/Abstract] OR "cerebell*"[Title/Abstract] OR "intracerebral"[Title/Abstract] OR "intracranial"[Title/Abstract] OR "subarachnoid"[Title/Abstract]) AND ("haemorrhage*"[Title/Abstract] OR "hemorrhage*"[Title/Abstract] OR "haematoma*"[Title/Abstract] OR "hematoma*"[Title/Abstract] OR "bleed*"[Title/Abstract])) OR ("hemipleg*"[Title/Abstract] OR "hemipar*"[Title/Abstract] OR "Paresis"[Title/Abstract] OR "paretic"[Title/Abstract])) AND ("Physical Therapy Modalities"[MeSH Terms:noexp] OR "Physical Therapy Specialty"[MeSH Terms:noexp] OR "Exercise Therapy"[MeSH Terms] OR "Hydrotherapy"[MeSH Terms:noexp] OR "Exercise Movement Techniques"[MeSH Terms] OR "Physical and Rehabilitation Medicine"[MeSH Terms:noexp] OR ("Rehabilitation"[MeSH Terms:noexp] OR "Recreation Therapy"[MeSH Terms:noexp]) OR ("Movement"[MeSH Terms:noexp] OR "Motor Activity"[MeSH Terms:noexp] OR "Exercise"[MeSH Terms:noexp] OR "Circuit-Based Exercise"[MeSH Terms:noexp] OR "physical conditioning, human"[MeSH Terms:noexp] OR "Plyometric Exercise"[MeSH Terms:noexp] OR "Resistance Training"[MeSH Terms:noexp] OR "Walking"[MeSH Terms:noexp] OR "Running"[MeSH Terms] OR "Swimming"[MeSH Terms:noexp]) OR "Sports for Persons with Disabilities"[MeSH Terms] OR ("Physical Exertion"[MeSH Terms:noexp] OR "Physical Endurance"[MeSH Terms] OR "Physical Fitness"[MeSH Terms:noexp]) OR ("Muscle Contraction"[MeSH Terms:noexp] OR "Isotonic Contraction"[MeSH Terms:noexp] OR "Isometric Contraction"[MeSH Terms:noexp]) OR ("physiotherap*"[Title/Abstract] OR ("physical"[Title/Abstract] AND ("mobilis*"[Title/Abstract] OR "mobiliz*"[Title/Abstract] OR "exercise*"[Title/Abstract] OR "exertion"[Title/Abstract] OR "endurance"[Title/Abstract] OR "therap*"[Title/Abstract] OR "conditioning"[Title/Abstract] OR "activit*"[Title/Abstract] OR "fitness"[Title/Abstract]))) OR ("Rehabilitation"[Title/Abstract] OR "Recovery of Function"[Title/Abstract] OR "exercise*"[Title/Abstract] OR "mobilis*"[Title/Abstract] OR "mobiliz*"[Title/Abstract] OR "motor activit*"[Title/Abstract] OR "motor skill*"[Title/Abstract]) OR ("Exercise"[Title/Abstract] AND ("train*"[Title/Abstract] OR "intervention*"[Title/Abstract] OR "protocol*"[Title/Abstract] OR "program*"[Title/Abstract] OR "therap*"[Title/Abstract] OR "activit*"[Title/Abstract] OR "regim*"[Title/Abstract])) OR ("fitness"[Title/Abstract] AND ("train*"[Title/Abstract] OR "intervention*"[Title/Abstract] OR "protocol*"[Title/Abstract] OR "program*"[Title/Abstract] OR "therap*"[Title/Abstract] OR "activit*"[Title/Abstract] OR "regim*"[Title/Abstract])) OR (("training"[Title/Abstract] OR "conditioning"[Title/Abstract]) AND ("intervention*"[Title/Abstract] OR "protocol*"[Title/Abstract] OR "program*"[Title/Abstract] OR "activit*"[Title/Abstract] OR "regim*"[Title/Abstract])) OR ("sport*"[Title/Abstract] OR "recreation*"[Title/Abstract] OR "leisure"[Title/Abstract] OR "cycling"[Title/Abstract] OR "bicycl*"[Title/Abstract] OR "rowing"[Title/Abstract] OR "treadmill*"[Title/Abstract] OR "Running"[Title/Abstract] OR "circuit training"[Title/Abstract] OR "swim*"[Title/Abstract] OR "walk*"[Title/Abstract] OR "dance*"[Title/Abstract] OR "dancing"[Title/Abstract] OR "tai ji"[Title/Abstract] OR "tai chi"[Title/Abstract] OR "yoga"[Title/Abstract]) OR (("endurance"[Title/Abstract] OR "aerobic"[Title/Abstract] OR "cardio*"[Title/Abstract]) AND ("fitness"[Title/Abstract] OR "train*"[Title/Abstract] OR "intervention*"[Title/Abstract] OR "protocol*"[Title/Abstract] OR "program*"[Title/Abstract] OR "therap*"[Title/Abstract] OR "activit*"[Title/Abstract] OR "regim*"[Title/Abstract])) OR ("muscle strengthening"[Title/Abstract] OR "progressive resist*"[Title/Abstract]) OR (("weight"[Title/Abstract] OR "strength*"[Title/Abstract] OR "resistance"[Title/Abstract]) AND ("train*"[Title/Abstract] OR "lift*"[Title/Abstract] OR "exercise*"[Title/Abstract])) OR (("isometric"[Title/Abstract] OR "isotonic"[Title/Abstract] OR "eccentric"[Title/Abstract] OR "concentric"[Title/Abstract]) AND ("action*"[Title/Abstract] OR "contraction*"[Title/Abstract] OR "exercise*"[Title/Abstract]))) AND ("physical activity"[Title/Abstract] OR "activit*"[Title/Abstract] OR "physical mobility"[Title/Abstract] OR "Exercise"[Title/Abstract] OR "Physical Exertion"[Title/Abstract] OR "Physical Endurance"[Title/Abstract] OR "energy expenditure"[Title/Abstract] OR "energy metabolism"[Title/Abstract] OR ("monitoring, physiologic"[MeSH Terms:noexp] OR "monitoring, ambulatory"[MeSH Terms] OR ("Fitness Trackers"[MeSH Terms:noexp] OR "Accelerometry"[MeSH Terms]) OR (("physical"[Title/Abstract] OR "physiolog*"[Title/Abstract] OR "perform*"[Title/Abstract] OR "fit"[Title/Abstract] OR "train*"[Title/Abstract] OR "activ*"[Title/Abstract] OR "endur*"[Title/Abstract] OR "Exercise"[Title/Abstract]) AND ("track*"[Title/Abstract] OR "monitor*"[Title/Abstract] OR "measur*"[Title/Abstract] OR "device*"[Title/Abstract] OR "app"[Title/Abstract])) OR (("step*"[Title/Abstract] OR "walk*"[Title/Abstract]) AND ("count*"[Title/Abstract] OR "meter*"[Title/Abstract] OR "daily"[Title/Abstract])) OR ("pedometer*"[Title/Abstract] OR "actigraph*"[Title/Abstract] OR "acceleromet*"[Title/Abstract]) OR (("cell*"[Title/Abstract] OR "smart*"[Title/Abstract] OR "mobile"[Title/Abstract] OR "android"[Title/Abstract] OR "internet"[Title/Abstract] OR "web"[Title/Abstract]) AND ("comput*"[Title/Abstract] OR "device"[Title/Abstract] OR "app"[Title/Abstract] OR "phone"[Title/Abstract]))))) AND (randomizedcontrolledtrial[Filter])

#### Search strategy on Excerpta Medica Database

| No. | Query | Results |
|---|---|---|
| #39 | #37 AND 'randomized controlled trial'/de AND 'article'/it | **3159** |
| #38 | #37 AND 'randomized controlled trial'/de | **4460** |
| #37 | #8 AND #25 AND #36 | **49348** |
| #36 | #26 OR #27 OR #28 OR #29 OR #30 OR #31 OR #32 OR #33 OR #34 OR #35 | **5717203** |
| #35 | pedometer*:ab,ti,kw OR actigraph*:ab,ti,kw OR acceleromet*:ab,ti,kw | **39544** |
| #34 | (step*:ab,ti,kw OR walk*:ab,ti,kw) AND near:ab,ti,kw AND (count*:ab,ti,kw OR meter*:ab,ti,kw OR daily:ab,ti,kw) | **1674** |
| #33 | ((physical OR physiolog* OR perform* OR fit* OR train* OR activ* OR endur* OR exercise) NEAR/3 (track* OR monitor* OR measur* OR device* OR app*)):ab,ti,kw | **636681** |
| #32 | 'accelerometer'/de OR 'accelerometry'/de OR 'actimetry'/de OR 'pedometer'/de | **33524** |
| #31 | ((cell* OR smart* OR mobile OR android OR internet OR web) NEAR/3 (comput* OR device OR app* OR phone)):ab,ti,kw | **218747** |
| #30 | 'telemedicine'/de OR 'telehealth'/de OR 'medical device'/de OR 'devices'/de | **543419** |
| #29 | 'computer'/de OR 'microcomputer'/de OR 'minicomputer'/de OR 'personal digital assistant'/de | **103542** |
| #28 | 'mobile phone'/exp OR 'smartphone'/de OR 'mobile application'/de | **48835** |
| #27 | 'monitor'/de OR 'ambulatory monitoring'/de OR 'self monitoring'/de OR 'personal monitor'/de OR 'personal monitoring'/de OR 'physiologic monitoring'/de | **36003** |
| #26 | 'physical activity':ab,ti,kw OR activit*:ab,ti,kw OR 'physical mobility':ab,ti,kw OR exercise:ab,ti,kw OR 'physical exertion':ab,ti,kw OR 'physical endurance':ab,ti,kw OR 'energy expenditure':ab,ti,kw OR 'energy metabolism':ab,ti,kw | **4533985** |
| #25 | #9 OR #10 OR #11 OR #12 OR #13 OR #14 OR #15 OR #16 OR #17 OR #18 OR #19OR #20 OR #21 OR #22 OR #23 OR #24 | **1963214** |
| #24 | 'muscle strengthening':ab,ti,kw OR 'progressive resist*':ab,ti,kw | **3442** |
| #23 | ((weight OR strength* OR resistance) NEAR/3 (train* OR lift* OR exercise*)):ab,ti,kw | **47062** |
| #22 | ((endurance OR aerobic OR cardio*) NEAR/3 (fitness OR train* OR intervention* OR protocol* OR program* OR therap* OR activit* OR regim*)):ab,ti,kw | **95826** |
| #21 | sport*:ab,ti,kw OR recreation*:ab,ti,kw OR leisure:ab,ti,kw OR cycling:ab,ti,kw OR bicycl*:ab,ti,kw OR rowing:ab,ti,kw OR treadmill*:ab,ti,kw OR running:ab,ti,kw OR 'circuit training':ab,ti,kw OR swim*:ab,ti,kw OR walk*:ab,ti,kw OR dance*:ab,ti,kw OR dancing:ab,ti,kw OR 'tai ji':ab,ti,kw OR 'tai chi':ab,ti,kw OR yoga:ab,ti,kw | **617351** |
| #20 | ((training OR conditioning) NEAR/3 (intervention* OR protocol* OR program* OR activit* OR regim*)):ab,ti,kw | **117342** |
| #19 | (fitness NEAR/3 (train* OR intervention* OR protocol* OR program* OR therap* OR activit* OR regim* OR centre* OR center*)):ab,ti,kw | **8851** |
| #18 | (exercise NEAR/3 (train* OR intervention* OR protocol* OR program* OR therap* OR activit* OR regim*)):ab,ti,kw | **90475** |
| #17 | rehabilitation:ab,ti,kw OR 'recovery of function':ab,ti,kw OR exercise*:ab,ti,kw OR mobilis*:ab,ti,kw OR mobiliz*:ab,ti,kw OR 'motor activit*':ab,ti,kw OR 'motor skill*':ab,ti,kw OR 'activities of daily living':ab,ti,kw OR adl:ab,ti,kw | **874412** |
| #16 | physiotherap*:ab,ti,kw OR ((physical NEAR/3 (mobilis* OR mobiliz* OR exercise* OR exertion OR endurance OR therap* OR conditioning OR activit* OR fitness)):ab,ti,kw) | **316684** |
| #15 | 'muscle contraction'/exp | **136286** |

| No. | Query | Results |
|---|---|---|
| #14 | 'sport'/de | **56588** |
| #13 | 'physical activity'/de OR 'climbing'/de OR 'cycling'/de OR 'jogging'/de OR 'running'/de OR 'swimming'/de OR 'walking'/exp OR 'weight lifting'/de | **376668** |
| #12 | 'muscle exercise'/exp | **15007** |
| #11 | ('rehabilitation'/de OR 'community based rehabilitation'/de OR 'constraint induced therapy'/de OR 'functional training'/de OR 'home rehabilitation'/de OR 'muscle training'/de OR 'neurorehabilitation'/exp) AND 'recreational therapy'/de OR 'rehabilitation care'/de OR 'telerehabilitation'/de OR 'vocational rehabilitation'/de | **30487** |
| #10 | 'exercise'/exp | **394268** |
| #9 | 'kinesiotherapy'/exp | **92083** |
| #8 | #1 OR #2 OR #3 OR #4 OR #5 OR #6 OR #7 | **1063735** |
| #7 | hempar*:ab,ti,kw OR hemipleg*:ab,ti,kw OR 'brain injur*':ab,ti,kw | **127972** |
| #6 | 'hemiparesis'/de OR 'hemiplegia'/de OR 'paresis'/de OR 'neurologic gait disorder'/de OR 'hemiplegic gait'/de | **57297** |
| #5 | ((brain* OR cerebr* OR cerebell* OR intracerebral OR intracran* OR subarachnoid) NEAR/5 (h?emorrhage* OR h?ematoma* OR bleed*)):ab,ti,kw | **30284** |
| #4 | ((brain* OR cerebr* OR cerebell* OR intracran* OR intracerebral) NEAR/5 (isch?emi*OR infarct* OR thrombo* OR emboli* OR occlus*)):ab,ti,kw | **114379** |
| #3 | stroke:ab,ti,kw OR poststroke:ab,ti,kw OR post-stroke:ab,ti,kw OR cerebrovasc*:ab,ti,kw OR 'brain vasc*':ab,ti,kw OR 'cerebral vasc*':ab,ti,kw OR cva*:ab,ti,kw OR apoplex*:ab,ti,kw OR sah:ab,ti,kw | **550491** |
| #2 | 'stroke patient'/de | **36057** |
| #1 | 'cerebrovascular disease'/de OR 'basal ganglion hemorrhage'/exp OR 'brain hematoma'/exp OR 'brain hemorrhage'/exp OR 'brain infarction'/exp OR 'brain ischemia'/exp OR 'carotid artery disease'/exp OR 'cerebral artery disease'/de OR 'cerebrovascular accident'/exp OR 'intracranial aneurysm'/exp OR 'occlusive cerebrovascular disease'/exp | **799642** |

#### Reason for exclusion

| Study | Reason for exclusion from the systematic review and meta-analysis |
|---|---|
| Aguiar et al., 2018 | Protocol study |
| Andersen et al., 2002 | Wrong intervention |
| Ardestani et al., 2019 | Wrong study design |
| Ashizawa et al., 2022 | Wrong setting |
| Ashizawa et al., 2021 | Wrong setting |
| Bjorkdahl et al., 2006 | Wrong outcomes |
| Boss et al., 2014 | Protocol study |
| Brauer et al., 2018 | Protocol study |
| Brouwer et al., 2018 | Wrong outcome |
| Chang et al., 2020 | Protocol study |
| Combs-Miller et al., 2014 | Wrong outcomes |
| Dean et al., 2021 | Protocol study |
| Dean et al., 2009 | Protocol Study |
| Dean et al., 2016 | Protocol Study |
| Dickstein et al., 2013 | Wrong intervention |
| Givon et al., 2015 | Wrong intervention |
| Gjelsvik et al., 2014 | Wrong population |
| Green et al., 2002 | Wrong intervention |
| Green et al., 2004 | Wrong study design |
| Gunnes et al., 2019 | Wrong study design |
| Hornby et al., 2008 | Wrong intervention |
| Hsieh et al., 2014 | Wrong outcomes |
| Johnson et al., 2018 | Protocol study |
| Katz-Leurer et al., 2003 | Wrong setting |
| Kirk et al., 2014 | Wrong population |
| Krarup et al., 2008 | Protocol study |
| Krawcyk et al., 2019 | Wrong population |
| Kwakkel et al., 2002 | Wrong study design |
| Langhammer et al., 2014 | Wrong outcomes |
| Lin et al., 2008 | Wrong outcomes |
| Lord et al., 2008 | Wrong outcomes |
| MacKay-Lyons et al., 2010 | Protocol study |
| Mansfield et al., 2017 | Protocol study |
| Mansfield et al., 2018 | Wrong comparator |
| Meester et al., 2019 | Wrong intervention |
| Mirelman et al., 2008 | Wrong intervention |
| Mura et al., 2022 | Protocol study |
| Olney et al., 2006 | Wrong intervention |

| | |
|---|---|
| Pastva et al., 2021 | Wrong study design |
| Plummer-D’Amato et al., 2012 | Protocol study |
| Pohl et al., 2007 | Wrong outcomes |
| Richardson et al., 2018 | Protocol study |
| Rooij et al., 2021 | Wrong intervention |
| Ryan et al., 2006 | Wrong intervention |
| SteenKrawcyk et al., 2019 | Protocol study |
| Teixeira-Salmela et al., 1999 | Wrong design |
| Timmermans et al., 2021 | Wrong outcomes |
| Torres-Arreola et al., 2009 | Wrong setting |
| VanderPloeg et al., 2006 | Wrong setting |
| Vanroy et al., 2019 | Wrong setting |
| Vluggen et al., 2021 | Wrong setting |
| Wade et al., 1992 | Wrong intervention |
| Walter et al., 2015 | Wrong study design |
| Wright et al., 2018 | Protocol study |
| Yang et al., 2008 | Wrong outcomes |
| Yeh et al., 2017 | Protocol study |
| Young et al., 1992 | Wrong intervention |
