## Supplemental M.3 for "Effectiveness of active physiotherapy on physical activity level in community-dwelling stroke survivors: a systematic review and meta-analysis of randomized controlled trials"

### Table S1: characteristics of included studies

<table>
<tr><td>Study Characteristics</td><td>Participants</td><td colspan="2">Intervention and exercise components</td><td colspan="2">Comparison</td><td>Outcome</td></tr>
<tr><td>Authors (Publication Yr)<br>(reference)<br><br>Country<br><br>Design</td><td>Sample Size n, Women (%)<br>Age Mean (SD) Yr<br>Delay since Stroke Yr (subacute/chronic phase)<br>Impairment Scale (Severity Classification)<br>Cognitive Function Scale (Level of Impairment)<br>BMI Mean (SD)<br>Balance Test and Score Mean (SD)<br>Gait Speed Scale, Score Mean (SD) and Classification</td><td>Brief description<br>Exercise Supervision<br>(Yes, No, or both, Who)<br>Group (Yes or No)<br>Setting<br>Program duration (wk)</td><td>Frequency<br>Intensity<br>Time (min)<br>Type</td><td>Description<br>Duration</td><td>Frequency<br>Intensity<br>Time (min)<br>Type</td><td>Objective or subjective measure<br>Tool<br>Primary or secondary outcome<br>Main findings<br>Post intervention follow-up (wk)</td></tr>
<tr><td>Aguiar et al. (2020)<br><br>Brazil<br><br>RCT, Parallel group</td><td>N= 22<br><br>IG: n=11, 27% Women<br>52(11) yr<br>4.25(5.67) yr since stroke: chronic phase<br>Fugl-Meyer Lower Limb: mild impairment<br>n.d.<br>26(3.3)<br>n.d,<br>10MWT,1.04(0.22) m/s, full community ambulator<br><br>CG: n=11, 27% Women<br>48 (10) yr<br>3.67 (2.17) yr since stroke: chronic phase<br>Fugl-Meyer Lower Limb: mild impairment<br>n.d.<br>29 (2.9)</td><td>Aerobic treadmill training at 60-80% of HRR (30 min with 5-min warm-up and 5-min cool-down)<br><br>Yes, physiotherapist<br><br>Yes, groups of 2-4 participants<br><br>n.d.<br><br>12 w.</td><td>F: 3 times per wk<br>I: vigorous<br>T: 40 min<br>T: cardiopulmonary intervention</td><td>outdoor-overground walking below 40% of HRR</td><td>F: 3 times per wk<br>I: low<br>T : 40 min<br>T : passive or less active intervention</td><td>Objective<br>Accelerometer<br>Primary<br>FU: 4 wk</td></tr>
</table>

| | | | | | | |
|---|---|---|---|---|---|---|
| | n.d.<br>10MWT, 0.97(0.19) m/s, full community ambulator | | | | | |
| Askim et al. (2018)<br><br><br>Norway<br><br>RCT, Parallel group | N= 380<br><br>IG: n= 186, 44%<br>71.7 (11.9) yr<br>0.3 (0.07) yr since stroke: subacute phase<br>Modified Rankin Scale: mild impairment<br>MMSE 27.8 (2.3): none<br>n.d.<br>TUG 12.3(7.77) s, independent for main transfers<br>6MWT,1.09(0.47) m/s, full community ambulator<br><br>CG: n= 194, 35%<br>72.3 (11.3) yr<br>0.31 (0.05) yr since stroke: subacute phase<br>Modified Rankin Scale: mild impairment<br>MMSE 27.9 (2.6): none<br>n.d.<br>TUG 16.1 (31.34) s, independent for main transfers<br>6MWT,1.08(0.65) m/s, full community ambulator | Schedule for physical activities and exercise set by a physiotherapist once a mth<br>+ Usual Care<br><br>No,<br>Physiotherapist<br><br>No,<br><br>Home,<br><br>78 w. | F: exercise = once a wk; physical activity = once a d<br>Usual care: once a wk<br>I: moderate to vigorous<br>Exercise: between 15 and 17 on Borg Scale<br>T: exercise = 45 to 60 min; physical activity = 30 min; usual care = 45 min<br>T : Multi component intervention | Usual Care<br>12 w. to 26 w. or longer depending on severity of disability | F: once a wk<br>I: moderate<br>T: 45 min<br>T: Usual Care | Subjective<br>IPAQ (MET. min/wk)<br>Secondary<br>No FU |
| Batchelor et al. (2012)<br><br><br>Australia<br><br>RCT, Parallel group | N= 156<br><br>IG: n= 71, 36.6%<br>70.8 (11.4) yr<br>0.25 (0.13) yr since stroke: subacute phase<br>Functional Independence Measure: mild impairment<br>n.d.<br>n.d.<br>n.d.<br>5MWT, 0.54 (0.27) m/s, limited community ambulator | OTAGO exercise program (home-based multifactorial falls prevention program)<br>+ Usual Care<br><br>No,<br>Physiotherapist<br><br>No,<br><br>Home, | F: exercise = 3 to 5 times per wk<br>Walking = 3 to 5 times per wk<br>I: low<br>T : exercise = 30 to 40 min<br>T : Multi component intervention | Usual Care (physiotherapy and occupational therapy) and falls prevention booklet | F: n.d.<br>I: sedentary<br>T: n.d.<br>T: Usual Care | Subjective<br>HAP-AAS<br>Secondary<br>No FU |

| | | | | | | |
|---|---|---|---|---|---|---|
| | CG: n= 85, 36.5%<br>72.2 (9.9) yr<br>0.26 (0.16) yr since stroke: subacute phase<br>Functional Independence Measure: mild impairment<br>n.d.<br>n.d.<br>n.d.<br>5MWT, 0.53 (0.33) m/s, limited community ambulator | 52w. | | | | |
| Boysen et al. (2009)<br><br>Denmark, China, Poland, Estonia<br><br>RCT, Parallel group | N= 314<br><br>IG: n= 157 (43.3%)<br>Modified Rankin Scale: No impairment<br>n.d.<br>25.8 (4.2)<br>n.d.<br>n.d.<br><br>CG: n= 157 (43.6%)<br>Modified Rankin Scale: No impairment<br>n.d.<br>25.9 (4.5)<br>n.d.<br>n.d. | Repeated encouragement and verbal instruction in a detailed training program (6 home visits and telephone calls)<br><br>No, Physiotherapist<br><br>No,<br><br>Home and community setting,<br><br>104w. | F: n.d.<br>I: low<br>T: n.d.<br>T : Multi component intervention | Usual Care + information on the possible benefits of physical activity but no specific instruction | F: n.d.<br>I: sedentary<br>T: n.d.<br>T: Usual Care | Subjective<br>PASE<br>Primary<br>No FU |
| Dean et al. (2012)<br><br>Australia<br><br>RCT, Parallel group | N=151<br><br>IG: n= 76 (50%)<br>66.7 (14.3) yr<br>6.7 (6.7) yr since stroke: chronic phase<br>n.d.<br>MMS, 27 (3): No cognitive impairment | WEEB (Weight-bearing Exercise for Better Balance) in a circuit-style group exercise class (40 sessions) + a home exercise program, + advice to increase walking | F: exercise class = once a wk<br>Home exercise program = 3 times per wk<br>I: moderate | exercise class to improve upper-limb function + home program (cognition) | F: exercise class = once a wk<br>Home exercise program = 3 times per wk<br>I: low | Objective<br>Number of steps per d for 7 consecutive ds (Digimax Pedometer)<br>Secondary |

| | | | | | | |
|---|---|---|---|---|---|---|
| | n.d.<br>TUG, 25 (28.3) m: requires assistance<br>10MWT, 0.72 (0.36) m/s: limited community ambulator<br><br>CG: n= 75 (47%)<br>67.5 (10.2) yr<br>5.2 (5.4) yr since stroke: chronic phase<br>n.d.<br>MMS, 27 (3): No cognitive impairment<br>n.d.<br>TUG, 30.2 (32.9) m: requires assistance<br>10MWT, 0.67 (0.38) m/s: limited community ambulator | Both, physiotherapist<br><br>Yes,<br><br>Home and community setting<br><br>52 w. | T: class and home programme = 45 to 60 min<br>T: multi-component intervention | | T: class and home programme = 45 to 60 min<br>T: passive or less active intervention | |
| Dean et al. (2018)<br><br>United Kingdom<br><br>RCT, Parallel group | N= 45<br><br>IG: n= 23 (30%)<br>70 (12) yr<br>3.86 (4.38) yr since stroke: chronic phase<br>Modified Rankin Scale: mild impairment<br>MMSE 27.5 (2.54): none<br>n.d.<br>TUG, 27.57 (27.57): requires assistance<br>n.d.<br><br>CG: n=22 (33%)<br>71 (10) yr<br>3.19 (2.81) yr since stroke: chronic phase<br>Modified Rankin Scale: mild impairment<br>MMSE 27.9 (3.01): none<br>n.d.<br>TUG, 21.24 (11.18): requires assistance | Re-Train: rehabilitation training (an introductory one-to-one session + 10 twice-wkly group classes + a closing one to one session + 3 drop-in sessions) + Usual Care<br><br>Yes, trainer<br><br>Yes, with up to two trainers and eight clients<br><br>Community setting<br><br>26 w. | F: twice a wk<br>I: moderate<br>T : 120 min<br>T= multi-component intervention | Usual Care + advice booklet about exercise after stroke | F: n.d.<br>I: low<br>T: n.d.<br>T: Usual Care | Objective<br>7-d physical activity level (GENEActiv, Accelerometer)<br>Primary<br>FU: 12 wk |

| | | | | | | |
|---|---|---|---|---|---|---|
| | n.d. | | | | | |
| Grau-Pellicer et al. (2020)<br><br>Spain<br><br>RCT, Parallel group | N= 41<br><br>IG: n= 24 (45.8%)<br>62.96 (11.87) yr<br>1.58 (2.3) yr since stroke: chronic phase<br>Barthel Index: mild impairment<br>n.d.<br>30.76(5.5)<br>TUG, 15.39 (6.37) m: independent for main transfers<br>10MWT, 0.82 (0.26) m/s: full community ambulator<br><br>CG: n=17 (52.9%)<br>68.53 (11.53) yr<br>1.74 (4.98) yr since stroke: chronic phase<br>Barthel Index: mild impairment<br>n.d.<br>30.14 (3.99)<br>TUG, 19.75 (9.17) m: independent for main transfers<br>10MWT, 0.57 (0.25) m/s: limited community ambulator | Multimodal Rehabilitation Program: 8 w supervised rehabilitation program with aerobic, task oriented. balance and stretching exercises + supervising adherence to physical activity through a mHealth app + ambulation program at home with the aim to reach 150 min/w of moderate physical activity<br><br>Yes, physiotherapist<br><br>Yes, 4-6 participants with 1 physiotherapist<br><br>Hospital or clinic<br><br>8 w. | F: twice a wk<br>I: moderate<br>T : 60 min<br>T : multi-component intervention | Conventional rehabilitation program: trunk exercise. muscle strengthening. OT. gait training<br><br>12 w. | F: n.d.<br>I: sedentary<br>T: n.d.<br>T: Usual Care | Subjective<br>Community ambulation: amount of outdoors walking time per d<br>Primary<br>No FU |
| Harrington et al. (2010)<br><br>United Kingdom<br><br>RCT, Parallel group | N= 243<br><br>IG: n= 119 (45%)<br>71(10.5) yr<br>n.d. chronic phase<br>Modified Barthel Index: mild impairment<br>MMSE, 27.67 (2.25): no impairment<br>n.d.<br>TUG, 20.0 (5.56) m: independent for main transfers<br>n.d. | Exercise session (circuit training (60 min) designed to improve balance, endurance, strength, flexibility, function, and well-being), followed by interactive education (60 min) session<br>+ home exercise manuals | F: twice a wk<br>I: moderate<br>T : 60 min<br>T : multi-component intervention | Usual Care | F: n.d.<br>I: n.d.<br>T: n.d.<br>T: Usual Care | Subjective<br>FAI<br>Primary<br>FU: 26 wk |

| | | | | | | |
|---|---|---|---|---|---|---|
| | CG: n= 124 (46%)<br>70 (10.2) yr<br>n.d. chronic phase<br>Modified Barthel Index: mild impairment<br>MMSE, 27.4 (2.85): no impairment<br>n.d.<br>TUG, 18.53 (3.38) m: independent for main transfers<br>n.d. | Yes, volunteers, and qualified exercise instructors supported by a physiotherapist<br><br>Yes, 1 instructor for 9 participants<br><br>Community setting<br><br>8 w. | | | | |
| Holmgren et al. (2010)<br><br>Sweden<br>RCT, Parallel group | N= 34<br><br>IG: n= 15 (40%)<br>77.7 (7.6) yr<br>0.38 (0.11) yr since stroke: subacute phase<br>Modified Rankin Scale: mild impairment<br>MMSE, 26.3 (3.5): mild impairment<br>n.d.<br>BBS, 44.3 (8.59): good balance<br>n.d.<br><br>CG: n= 19 (37%)<br>79.2 (7.5)<br>0.35 (0.08) yr since stroke: subacute phase<br>Modified Rankin Scale: mild impairment<br>MMSE, 25.5 (4.4): mild impairment<br>BBS, 44.2 (11.01): good balance<br>n.d. | HIFE (High Intensity Functional exercise) program + activities related to real-life situations) + education on falls and security aspects (once a wk)<br><br>Yes, physiotherapist and occupational therapist<br><br>Yes,<br><br>Hospital or clinic<br>5w.<br><br>+ an individualized home-based exercise program (based on exercises performed during the 5 | F: 6 times per wk<br>I: High<br>T: 90 min (45 min HIFE session + 30 min break+ 45 min activities session)<br>T : multi-component intervention | Group discussions about hidden dysfunctions after stroke | F: once a wk<br>I: sedentary<br>T: 60 min<br>T: Passive or less active intervention<br><br>5w. | Subjective<br>FAI-3<br>Secondary<br>FU: 12 wk |

| | | | | | | |
|---|---|---|---|---|---|---|
| | | wk) to perform 3 times per wk until the 3-mth FU | | | | |
| Ivey et al. (2015)<br><br>USA<br><br>RCT, Parallel group | N= 34<br>IG: n=18 (44%)<br>61 (6.79) yr<br>3.42 (4.24) yr since stroke: chronic phase<br>n.d.<br>n.d.<br>25.7 (4.24)<br>n.d.<br>6MWT, 0.66 (0.38) m/s : limited community ambulator<br><br>CG: n=16 (31%)<br>63 (9.6) yr<br>3.08 (4.67) yr since stroke: chronic phase<br>n.d.<br>n.d.<br>26.9 (4.8)<br>n.d.<br>6MWT, 0.48 (0.24) m/s : limited community ambulator | Higher intensity treadmill training (progressive training to a target intensity of 80% HRR)<br><br>Yes, trainer<br><br>No,<br><br>Laboratory<br><br>26 w. | F: twice a wk<br>I: high<br>T: 30 min<br>T: cardiopulmonary intervention | Lower intensity treadmill training (training intensity of less than 50% HRR), with higher duration to maintain comparable caloric expenditure between groups | F: twice a wk<br>I: moderate<br>T: n.d.<br>T: Passive or less active intervention | Objective<br>48-hour ambulatory activity (Step Activity Monitor technology)<br>Secondary<br>No FU |
| Kono et al. (2013)<br><br>Japan,<br><br>RCT, Parallel group | N= 70<br><br>IG: n= 35 (40%)<br>63.5 (7.0) yr<br>n.d. subacute phase<br>n.d.<br>n.d.<br>23.1 (2.19) | Lifestyle intervention (exercise training + salt restriction + nutrition advice)<br>Exercise training = home-exercise (instruction about increasing the amount of daily physical activity + | F: Lifestyle intervention= once or twice per wk<br>Walking prescription= 3-5 per wk<br>Home exercise = 2 sessions | Advice + usual medical care<br><br>24 w. | F: 3 sessions in 24 w.<br>I: sedentary<br>T: n.d.<br>T: Passive or less active intervention | Objective<br>Mean daily step count Accelerometer 24h/d for 1 wk (Kenz Lifecorder, Suzuken, Nagoya, Japan)<br>Secondary<br>No FU |

| | | | | | | |
|---|---|---|---|---|---|---|
| | n.d.<br>n.d.<br><br>CG: n= 35 (22.9%)<br>63.4 (11.4) yr<br>n.d. subacute phase<br>n.d.<br>n.d.<br>23.1 (2.74)<br>n.d.<br>n.d. | prescription of walking exercise) and center-based exercise training (aerobic exercise using a cycle ergometer + resistance training)<br>Both, physiotherapist<br>No,<br>Laboratory<br>24 wk | I: center-based exercise = moderate<br>T: center-based exercise = 60 min<br>Walking prescription = 30 to 60 min<br>T: Multi-component intervention | | | |
| Lennon et al. (2008)<br><br>Ireland,<br><br>RCT, Parallel group | N= 48<br>IG: n= 24 (42%)<br>59.0 (10.3) yr<br>4.56 (2.13) yr since stroke: chronic phase<br>n.d.<br>n.d.<br>28.1 (5.6)<br>n.d.<br>n.d.<br>CG: n= 24 (42%)<br>60.5 (10.0) yr<br>4.72 (3.27) yr since stroke: chronic phase<br>n.d.<br>n.d.<br>25.7 (3.7)<br>n.d.<br>n.d. | Cardiac rehabilitation program with cycle ergometry using either the upper or lower limbs (16 sessions) at 50 to 60% of HRMax and stress management classes (2 sessions)<br>+ Usual Care<br><br>Yes, physiotherapist,<br>No,<br><br>Hospital or clinic<br><br>10w. | F: twice a wk<br>I: moderate<br>T: 30 min<br>T: Cardiopulmonary intervention | Usual Care (PT + OT) excluding aerobic exercise | F: n.d.<br>I: low<br>T: n.d.<br>T: n.d. | Subjective<br>FAI<br>Secondary<br>No FU |
| Lin-Rong Liao et al. (2016) | N= 84<br><br>IG1: n=28 (29%) | Exercises on a WBV platform with same | F: 3 times a wk<br>I: IG1: low, IG2: high | Exercises on the WBV | F: 3 times a wk<br>I: IG1: low, IG2: low | Subjective<br>FAI<br>Secondary |

| | | | | | | |
|---|---|---|---|---|---|---|
| China<br><br>RCT, Parallel group | 60.8 (8.3) yr<br>8.5 (5.2) yr since stroke<br>n.d.<br>n.d.<br>24.1 (5.8) kg.m-2<br>TUG, 20.2 (14.8) m: requires assistance<br>6MWT, 0.57 (0.23) m/s: limited community ambulator<br><br>IG2: n= 28 (36%)<br>62.9 (10.2) yr<br>8.1 (4.2) yr since stroke, chronic phase<br>n.d.<br>n.d.<br>24.7 (3.2) kg.m-2<br>TUG, 17.9 (9) m: independent for main transfers<br>6MWT, 0.58 (0.21) m/s: limited community ambulator<br><br>CG: n=28 (14%)<br>59.8 (9.1) yr<br>9.0 (4.6) yr since stroke, chronic phase<br>n.d.<br>n.d.<br>25.1 (3.8) kg.m-2<br>TUG, 22.4 (24) m: requires assistance<br>6MWT, 0.58 (0.21): limited community ambulator | dynamic and static exercises<br>IG1: Low intensity Whole Body Vibration, LWBV (20Hz, 1mm)<br>IG2: High intensity Whole Body Vibration, HWBV (30Hz, 1 mm)<br><br>Yes, trainer<br><br>Yes, ratio 1 trainer and 2 participants<br><br>Laboratory,<br><br>12 w. | T: IG1: 12 min, IG2: 18 min<br>T: Musculo-skeletal intervention | platform turned off<br><br>12w. | T: IG1: 12 min, IG2: 18 min<br>T: Passive or less active intervention | No FU |
| Martins et al. (2020)<br><br>Brazil | N= 36<br><br>IG: n=18 (56%)<br>56 (17) yr | task-specific training: organized in a circuit with 11-station that included activities of reaching, | F: 3 times a wk<br>I: moderate<br>T: 60 min (30 min of tasks for the upper | global stretching, memory exercises, and | F: 3 times a wk<br>I: low<br>T: 60 min | Objective<br>Total daily energy expenditure (kilojoules), Activity monitor SenseWear multisensory. |

| | | | | | | |
|---|---|---|---|---|---|---|
| RCT, Parallel group | 4.33 (4.29) yr since stroke: chronic phase<br>Fugl-Meyer LL, mild impairment<br>n.d.<br>27 (7) kg.m-2<br>n.d.<br>6MWT, 0.94 (0.34) m/full community ambulator<br><br>CG: n=18 56%)<br>55(13) yr<br>3.42 (2.6) yr since stroke: chronic phase<br>Fugl-Meyer LL, mild impairment<br>n.d.<br>26 (5) kg.m-2<br>n.d.<br>6MWT, 0.95 (0.38): full community ambulator | grasping, manipulation of different objects, writing, sit-to-stand, step, and heel raise activities, and walking<br><br>Yes, physiotherapist<br><br>Yes, in groups of two to six<br><br>heath centers and laboratory settings<br><br>12 w. (36 sessions) | limbs and 30 min of tasks for the lower limb) (five min of exercises in each station, except for the gait training, which lasted 10 min)<br>T: functional task training | health education sessions (Most of the stretches were performed in a sitting or lying | T: passive or less active intervention | Primary<br>FU: 4 wk |
| Mayo et al. (2015)<br><br>Canada<br><br>RCT, Cross-over | N= 186<br><br>IG: n= 93 (39%)<br>61 (12) yr<br>2.5 (2.2) yr since stroke: chronic phase<br>n.d.<br>n.d.<br>n.d.<br>n.d., 0.82 (0.33) m/s: full community ambulator<br>CG: n= 93 (40%)<br>65 (11) yr<br>3.1 (3.1) yr since stroke: chronic phase<br>n.d.<br>n.d.<br>n.d.<br>n.d. | The Getting on with the Rest of Your Life: Mission Possible © (home-based intervention included exercise and project-based activities promoting learning, leisure, and social activities), exercise component is based on FAME program with Five key elements were incorporated: aerobic exercise, strength of peripheral and core musculature, balance, | F: twice a wk<br>I: moderate<br>T : 45 min<br>T : multi-component intervention | Wait list (12 w.) | F: n.d.<br>I: sedentary<br>T: n.d.<br>T : no treatment | Subjective<br>Time spent in meaningful activity outside of the home excluding activities directly provided by the program with CHAMPS Community Healthy Activities Model Program for Seniors) questionnaire<br>Primary |

| | | | | | | |
|---|---|---|---|---|---|---|
| | n.d., 0.76 (0.42) m/s: limited community ambulator | flexibility, and rapidity of movements.<br><br>yes, recreation therapists. educators. exercise therapists or other personnel with experience in healthcare and with stroke<br><br>Yes,<br><br>Community setting<br><br>52 w. | | | | |
| Moore et al. (2010)<br><br>USA<br><br>RCT, Crossover | N= 20 (30%)<br>Number of participants in IG and CG: n.d.<br>50 (15) yr<br>1.08 (0.67) yr since stroke: chronic phase<br>n.d.<br>n.d.<br>n.d.<br>TUG, 23(11) m : independent for main transfers<br>n.d., 0.51 (0.21) m/s: limited community ambulator<br><br>IG : n= n.d. (30%)<br>CG : n=n.d (30%) | Body weight supported treadmill training<br><br>Yes, n.d.<br><br>No,<br><br>n.d.<br><br>4w. | F: 2 to 5 per wk<br>I: high<br>T:<br>T: Cardiopulmonary intervention | Delayed locomotor training group | F: n.d.<br>I: sedentary<br>T: n.d.<br>T: No treatment | Objective<br>Daily stepping activity Step Activity Monitor<br>Secondary<br>No FU |
| Mudge et al. (2009)<br><br>New Zealand<br>RCT, Parallel group | N= 58<br><br>IG: n= 31 (39%) | Lower-limb circuit exercise-based rehabilitation: 15 stations containing either a task- | F: 3 times a wk<br>I: moderate<br>T: exercise: 30 min (2 min at each station) | 4 social and 4 educational sessions, matched dose | F: 2 times per wk<br>I: low<br>T: 90 min | Objective<br>Steps/d (3 ds) StepWatch Activity Monitor<br>Primary |

| | | | | | | |
|---|---|---|---|---|---|---|
| | 70 (12.5) yr<br>5.14 (3.18) yr since stroke: chronic phase<br>Rivermead Mobility Index, mild impairment<br>n.d.<br>n.d.<br>n.d.<br>10MWT, 0.76 (0.3) m/s: limited community ambulator<br>CG: n=27 (52%)<br>68 (10.5) yr<br>7.7 (4.55) yr since stroke: chronic phase<br>Rivermead Mobility Index, mild impairment<br>n.d.<br>n.d.<br>n.d.<br>10MWT, 0.62 (0.27) m/s: limited community ambulator | oriented gait or standing balance activity, or strengthening<br><br>Yes, physiotherapist and 2 physiotherapist students<br><br>Yes (The groups contained up to 9 participants and were led by 1 of the investigators assisted by 2 physiotherapy students)<br><br>Private rehabilitation clinic<br><br>4 w. | Stretching: 20 to 30 min<br>T : multi component intervention | occupational therapist,<br><br>in groups of up to 8<br><br>4 w. | T: passive or less active intervention | FU: 12 wk |
| Pang et al. (2005)<br><br>Canada<br>RCT, Parallel group | N= 63<br><br>IG: n= 32 (41%)<br>65.8 (9.1) yr<br>5.2 (5) yr since stroke: chronic phase<br>n.d.<br>MMSE, 27.6 (2.3): No cognitive impairment<br>n.d.<br>BBS, 47.6(6.7): good balance<br>6MWT, 0.91 (0.4) m/s: full community ambulator<br><br>CG: n= 31 (42%)<br>64.7 (8.4) yr<br>5.1 (3.6) yr since stroke: chronic phase<br>n.d. | FAME, Fitness, and mobility exercise program, designed to improve cardiorespiratory fitness, mobility, leg muscle strength, balance, and hip bone mineral density (walking and stepping)<br><br>Yes, physiotherapist, occupational therapist, and exercise instructor, | F: 3 sessions per wk<br>I: moderate<br>T : 60 min<br>T : multi-component intervention | Seated upper extremity program | F: 3 times per w.<br>I: low<br>T: 60 min<br>T: passive or less active intervention | Subjective<br>PASIPD<br>Primary<br>No FU |

| | | | | | | |
|---|---|---|---|---|---|---|
| | MMSE, 28.2 (1.9): No cognitive impairment<br>n.d.<br>BBS, 47.3 (6.1): good balance<br>6MWT, 0.84 (0.34) m/s: full community ambulator | Yes, 9 to 12 participants supervised by the 3 providers<br><br>Community setting<br><br>19 w. | | | | |
| Pang et al. (2018)<br><br>China<br>RCT, Parallel group | N=84<br><br>IG 1: n= 28 (21%)<br>59.9 (6.8) yr<br>6 (5.3) yr since stroke: chronic phase<br>n.d.<br>MoCA, 25.9 (2.7): no cognitive impairment<br>23.7 (2.7) kg.m-2<br>Mini-BEST Test, 16.6 (5.2): high risk for fall<br>n.d.<br><br>IG 2: n= 28 (29%)<br>61.2 (6.2) yr<br>5.55 (3.47) yr since stroke: chronic phase<br>n.d.<br>MoCA, 25.6 (2.6): no cognitive impairment<br>23.5 (3.2) kg.m-2<br>Mini-BEST Test, 18.1 (4.4)<br>n.d.<br><br>CG: n=28 (36%)<br>62.4 (6.3) yr<br>7.29 (6.94) yr since stroke: chronic phase<br>n.d.<br>MoCA, 26.4 (2.9): no cognitive impairment | 3 arms<br>IG1: Dual Task balance/mobility training<br>IG2: Single Task balance/mobility training<br><br>Yes, Physiotherapist,<br><br>Yes, group with a ratio of 2 instructors for 4 participants<br><br>Community-setting (exercise room located in the university)<br><br>8 w. | F: 3 times per wk<br>I: low<br>T: 60 min<br>T: functional task training | Sham exercise (flexibility exercises of all limbs and strengthening exercises of the upper limbs without additional cognitive tasks. The exercises were performed mainly in a sitting or lying position) | F: 3 times per wk<br>I: low<br>T: 60 min<br>T: passive or less active intervention | Subjective<br>FAI<br>Secondary<br>FU: 26 wk |

| | | | | | | |
|---|---|---|---|---|---|---|
| | 23.7 (3.9) kg.m-2<br>Mini-BEST Test, 17.4 (5.1): high risk for falls<br>n.d. | | | | | |
| Severinsen et al. (2014)<br><br><br>Denmark<br>RCT, Parallel group | N= 43<br>IG1: n=13 (31%)<br>IG2: n= 14 (21%)<br>CG: n= 16 (31%)<br>66 (22.76) yr<br>1.75 (1.96) yr since stroke: chronic phase<br>n.d.<br>n.d.<br>29.22 (12.4) kg.m-2<br>n.d.<br>10MWT, 0.78 (0.81) m/s: limited community ambulator | 3 arms<br>IG1: High-intensity Aerobic Training (cycle ergometer)<br>IG2: progressive Resistance Training<br><br>Yes, physiotherapist,<br>n.d.<br><br>12 w. | F: 3 sessions per wk<br>I: vigorous<br>T: 60 min<br>T: IG1: cardiopulmonary intervention<br>IG2: Musculo skeletal intervention | Low-intensity sham training of the arms | F: 3 times per wk<br>I: low<br>T: 60 min<br>T: Sham | Subjective<br>Physical Activity Scale (PAS) scores in metabolic equivalents<br>Secondary<br>FU: 52 wk |
| Shaughnessy et al. (2012)<br><br>USA<br><br>RCT, Parallel group | N= 113<br>IG: n= 57<br>n.d.<br>n.d. chronic phase<br>n.d.<br>n.d.<br>n.d.<br>n.d.<br>n.d.<br><br><br>CG: n= 56<br>n.d.<br>n.d. chronic phase<br>n.d.<br>n.d. | Task-oriented treadmill training program focused on the goal of engaging in exercise at an aerobic intensity of 60% HRR.<br>Yes, physiotherapist<br><br>No,<br><br>Community setting<br><br>26 w. | F: 3 times per wk<br>I: moderate<br>T: 40 min<br>T: cardiopulmonary intervention | Exercise-based treatment focused on stretching program (13 sessions)<br><br><br>26 w. | F: 3 times per wk<br>I: low<br>T: 40 min<br>T: passive or less active intervention | Subjective<br>YPAS, Yale Physical Activity Survey<br>primary<br>No FU |

| | | | | | | |
|---|---|---|---|---|---|---|
| | n.d.<br>n.d.<br>n.d. | | | | | |
| Shim et al. (2015)<br><br><br>Korea<br>RCT, Parallel group | N= 20<br><br>IG: n= 10 (10%)<br>54.3 (18.5) yr<br>0.66 (0.17) yr since stroke: chronic phase<br>FIM, moderate impairment<br>MMSE, 22.7 (4.5): mild cognitive impairment<br>n.d.<br>n.d.<br>n.d.<br><br>CG: n=10 (20%)<br>58.7 (9.9) yr<br>0.64 (0.18) yr since stroke: chronic phase<br>FIM, moderate impairment<br>MMSE, 17.8 (8.1): moderate cognitive impairment<br>n.d.<br>n.d.<br>n.d. | Bilateral arm training: functional tasks with both hands symmetrically (e.g., table cleaning, opening, and closing drawers, moving objects)<br><br>Yes, physiotherapist<br><br>No,<br><br>n.d.<br><br>6w. | F : 5 times per wk<br>I : low<br>T : 30 min<br>T : Functional task training | Unilateral arm training: functional task with only the affected hand (e.g., table cleaning, opening, and closing drawers, moving objects) | F: 5 times per wk<br>I: low<br>T : 30 min<br>T : passive or less active intervention | Objective<br>Amount of physical activity of the arm, triaxial accelerometer (Actisleep, GT3X+, LLT, USA)<br>Primary<br>No FU |
| Towfighi et al. (2020)<br><br><br>USA<br><br>RCT, Parallel group | N= 100<br><br>IG : n= 49 (41%)<br>60 (7) yr<br>n.d. subacute phase<br>n.d.<br>n.d.<br>29.7 (5.1) kg.m-2 | HEALS intervention: Healthy Eating and Lifestyle After Stroke (didactic presentations on a specific lifestyle practice + peer exchange + personal exploration + direct experience through | F: once a wk (6 sessions)<br>I : n.d.<br>T : 120 min (exercise time n.d.)<br>T : multi-component intervention | Usual Care | F : n.d.<br>I : n.d.<br>T : n.d.<br>T : n.d. | Subjective<br>Number of participants inactive, active, or doing health enhancing physical activity<br>Primary<br>FU: 26 wk |

| | | | | | | |
|---|---|---|---|---|---|---|
| | n.d.<br>n.d.<br><br>CG: n=51 (35%)<br>57 (10) yr<br>n.d. subacute phase<br>n.d.<br>n.d.<br>29.9 (8.9) kg.m-2<br>n.d.<br>n.d. | participation in activities (e.g., preparing meals, walking at a park, strengthening, stretching and yoga)<br><br>Yes, occupational therapist<br><br>Yes, 2 to 10 participants<br><br>Outpatient clinics<br>6 w. | | | | |
| Vahlberg et al. (2017)<br><br>Sweden<br>RCT, Parallel group | N= 67<br><br>IG : n= 34 (20.6%)<br>72.6 (5.5) yr<br>1.08 (0.33) yr since stroke: chronic phase<br>n.d.<br>n.d.<br>n.d.<br>BBS, 47.9 (9.6): good balance<br>10MWT, 1 (0.35) m/s: full community ambulator<br>CG: n= 33 (27.3%)<br>73.7 (5.3) yr<br>1.08 (0.17) yr since stroke: chronic phase<br>n.d.<br>n.d<br>n.d.<br>BBS, 49 (9.7): good balance<br>10MWT,1.08 (0.3) m/s: full community ambulator | Circuit classes. consisting of different workstations with functional exercises that involved the major muscle groups (particularly lower extremity function). Exercises are retrieved from the HIFE program<br><br>Yes, physiotherapist and assistant<br><br>Yes, group, ratio of the trainers: one physiotherapist and one assistant and 2 to 7 participants | F: twice wkly<br>I: low to moderate<br>T: warm-up with stationary cycling or walking (10 min) circuit class (45 min) motivational session (20 min)<br>T : multi-component intervention | Ordinary physical activities and rehabilitation programs | F: n.d.<br>I: low<br>T:n.d.<br>T : passive or less active intervention | Subjective<br>PASE<br>Secondary<br>FU : 12wk, 48wk |

| | | | | | | |
|---|---|---|---|---|---|---|
| | | 12w. | | | | |
| Wright et al. (2020)<br><br><br>United Kingdom<br><br>RCT, Parallel group | N=34<br><br>IG: n= 16 (12%)<br>59.6 (10.1) yr<br>2.58 (1.58) yr since stroke: chronic phase<br>Modified Rankin Scale, moderate impairment<br>n.d.<br>n.d.<br>TUG, 36.2 (20.2) m: requires assistance<br>6MWT, 0.38 (0.23) m/s: home ambulator<br>CG: n= 18 (22%)<br>65.1 (10.1) yr<br>2.67 (1.75) yr since stroke: chronic phase<br>Modified Rankin Scale, moderate impairment<br>n.d.<br>n.d.<br>TUG, 36 (21.6) m: requires assistance<br>6MWT, 0.34 (0.26) m/s: home ambulator | O-RAGT: Over-Ground Robotic-Assisted Gait Training program,<br>+ Usual Care (physiotherapy)<br><br>No, physiotherapist<br>No,<br><br><br><br>10wk | F: 7 times per wk<br>I: moderate<br>T: ≥ 30 min<br>T: functional task training | Physical Activity<br>+ Usual Care (physiotherapy)<br><br>10 w. | F: Physical activity = 7 times per wk<br>I: low<br>T: physical activity = 30 min<br>T: passive or less active intervention | Objective<br>Number of steps per d using accelerometry (ActivPAL3 device) 7 consecutive ds and nights<br>Secondary<br>FU: 22 wk |

CG = Control Group, IG = Intervention Group, n.d. = non determined, BMI = Body Mass Index, HRR = Heart Rate Reserve, TUG = Timed Up and Go, 10MWT = 10 Meter Walk Test, 6MWT = 6 Meter Walk Test, MMSE = Mini-Mental State Examination, BBS = Berg Balance Scale, FIM = Functional Independence Measure, MoCA = Montreal Cognitive Assessment, Mini BEST Test = Mini Balance Evaluation System Test, LL = Lower Limb, UL = Upper L
