## Supplemental M.4 for "Effectiveness of active physiotherapy on physical activity level in community-dwelling stroke survivors: a systematic review and meta-analysis of randomized controlled trials"

### Table S2: Meta-analysis of standardized mean differences

| Analysis | Standardized mean differences (95%CI) | N studies | N sample | Heterogeneity | | P value difference |
|---|---|---|---|---|---|---|
| | | | | I², % | P value | |
| **Overall** | 0.22 (0.04; 0.40) | 23 | 1834 | 65.4 | < 0.0001 | - |
| - Low risk of bias | 0.15 (0.03; 0.27) | 9 | 1011 | 17.4 | 0.288 | - |
| **Physical activity measurement** | | | | | | |
| - Objective | 0.48 (0.03; 0.92) | 9 | 424 | 72.8 | 0.003 | 0.081 |
| - Subjective | 0.06 (-0.08; 0.21) | 14 | 1410 | 34.9 | 0.096 | |
| **Types of comparators** | | | | | | 0.029 |
| - Passive or less active | 0.34 (0.10;0.57) | 16 | 826 | 61.8 | 0.0006 | |
| - Usual Care | -0.01 (-0.19;0.19) | 7 | 1008 | 50.4 | 0.06 | |
| **Mean age** | | | | | | |
| - < 65 yr | 0.39 (0.10; 0.68) | 13 | 558 | 62.8 | 0.001 | 0.063 |
| - ≥ 65 yr | 0.06 (-0.13; 0.25) | 10 | 1276 | 57.7 | 0.011 | |
| **Mean time since stroke** | | | | | | |
| - < 3 yr | 0.23 (0.02; 0.44) | 7 | 578 | 11.8 | 0.340 | 0.898 |
| - ≥ 3 yr | 0.21 (-0.09; 0.51) | 12 | 627 | 65.5 | 0.0008 | |
| **Phase** | | | | | | |
| - Subacute | 0.24 (-0.08; 0.56) | 5 | 772 | 67.2 | 0.016 | 0.594 |
| - Chronic | 0.21 (-0.01; 0.43) | 18 | 1062 | 65.6 | < 0.0001 | |
| **Impairment severity** | | | | | | |
| - None | 0.08 (-0.16; 0.31) | 1 | 276 | NA | NA | 0.208 |
| - Mild | 0.05 (-0.20; 0.30) | 9 | 828 | 57.7 | 0.015 | |
| - Moderate | 0.59 (0.03; 1.15) | 2 | 51 | 0.0 | 0.722 | |
| **Balance classification** | | | | | | |
| - Require assistance | 0.19 (-0.19; 0.58) | 3 | 194 | 38.2 | 0.186 | 0.704 |
| - Independent for main transfers | -0.01 (-0.28; 0.26) | 5 | 629 | 57.8 | 0.046 | |
| - Good balance | 0.07 (-0.39; 0.53) | 3 | 164 | 53.9 | 0.108 | |
| **Gait classification** | | | | | | |
| - Full community ambulator | 0.36 (-0.40; 1.13) | 5 | 409 | 81.7 | 0.0002 | 0.653 |
| - Home ambulator | 0.51 (-0.20; 1.23) | 1 | 31 | NA | NA | |
| - Limited community ambulator | 0.19 (-0.05; 0.42) | 7 | 525 | 44.2 | 0.10 | |
| **Types of intervention** | | | | | | |
| - Cardiopulmonary | 0.39 (-0.63; 1.41) | 4 | 160 | 84.6 | 0.002 | 0.506 |
| - Musculoskeletal | 0.03 (-0.34; 0.40) | 2 | 112 | 0.0 | 0.742 | |
| - Functional task-training | 0.38 (0.08; 0.67) | 5 | 182 | 0.0 | 0.684 | |
| - Multi-component | 0.17 (-0.05; 0.40) | 12 | 1380 | 70.7 | 0.001 | |
| **Supervision** | | | | | | |
| - yes | 0.20 (-0.04; 0.44) | 17 | 939 | 67.2 | < 0.0001 | 0.244 |
| - no | 0.08 (-0.07; 0.23) | 4 | 699 | 0 | 0.64 | |
| - both | 0.62 (-0.04; 1.28) | 2 | 196 | 78.8 | 0.03 | |
| **Supervision** | | | | | | |
| - Trainer | 0.40 (-0.57; 1.37) | 4 | 181 | 84.9 | 0.0002 | 0.42 |
| - Physio | 0.18 (0.02; 0.34) | 19 | 1720 | 84.1 | 0.001 | |
| **Frequency** | | | | | | |
| - 2 or 3 per wk | 0.50 (-0.23; 1.21) | 5 | 402 | 88.0 | < 0.0001 | 0.495 |
| - 4 to 7 per wk | 0.24 (0.05; 0.43) | 15 | 859 | 41.0 | 0.040 | |

| Analysis | Standardized mean differences (95%CI) | N studies | N sample | Heterogeneity | | P value difference |
|---|---|---|---|---|---|---|
| | | | | $I^2$, % | P value | |
| **Duration of intervention** | | | | | | |
| - 0-12 wk | 0.20 (-0.01; 0.41) | 14 | 778 | 51.8 | 0.012 | 0.965 |
| - 13-24 wk | 0.31 (-0.97; 1.60) | 2 | 133 | 92.5 | 0.0002 | |
| - > 24 wk | 0.25 (-0.16; 0.66) | 7 | 923 | 72.4 | 0.001 | |
| **Intensity** | | | | | | |
| - Vigorous | 0.30 (-0.20; 0.81) | 7 | 495 | 72.7 | 0.0012 | 0.829 |
| - Moderate | 0.20 (-0.10; 0.50) | 10 | 752 | 75.8 | < 0.0001 | |
| - Low | 0.15 (-0.01; 0.31) | 6 | 587 | 0.4 | 0.507 | |

NA = not applicable; CI = Confidence Intervals

### <u>Table S3</u>: Meta-regression

| **Variables** | **Univariable models** | | | |
|---|---|---|---|---|
| | **Coeff (95%CI)** | **P value** | **P value, test of moderators** | **$R^2$, %** |
| **Physical activity measurement** | | | 0.025 | 35.29 |
| - Objective | ref | | | |
| - Subjective | -0.386 (-0.724; -0.048) | 0.025 | | |
| **Types of comparator** | | | 0.071 | 23.24 |
| - Passive or less active intervention | ref | | | |
| - Usual Care | -0.3148 (-0.6561;0.0266) | 0.071 | | |
| **Mean age** | | | 0.047 | 22.5 |
| - < 65 yr | ref | | | |
| - ≥ 65 yr | -0.322 (-0.640; -0.004) | 0.047 | | |
| **% Females, increase of 1%** | -0.755 (-2.487; 0.977) | 0.393 | 0.393 | 0.6 |
| **Mean time since stroke** | | | 0.607 | 0.0 |
| - < 3 yr | ref | | | |
| - ≥ 3 yr | -0.103 (-0.493; 0.288) | 0.607 | | |
| **Phase** | | | 0.881 | 0.0 |
| - Chronic | ref | | | |
| - Subacute | 0.032 (-0.389; 0.453) | 0.881 | | |
| **Impairment severity** | | | 0.309 | 0.0 |
| - Mild | ref | | | |
| - None | 0.033 (-0.570; 0.636) | 0.015 | | |
| - Moderate | 0.554 (-0.156; 1.264) | 0.103 | | |
| **Balance classification** | | | 0.309 | 0.0 |
| - Good balance | ref | | | |
| - Require assistance | 0.122 (-0.475; 0.719) | 0.689 | | |
| - Independent for main transfers | -0.066 (-0.579; 0.446) | 0.800 | | |
| **Gait classification** | | | 0.832 | 0.0 |
| - Full community ambulator | ref | | | |
| - Home ambulator | 0.183 (-1.051; 1.417) | 0.771 | | |
| - Limited community ambulator | -0.131 (-0.748; 0.485) | 0.677 | | |
| **Types of intervention** | | | 0.760 | 0.0 |
| - Cardiopulmonary | ref | | | |
| - Musculoskeletal | -0.300 (-1.106; 0.506) | 0.466 | | |

| | | | | |
|---|---|---|---|---|
| - Functional task-training | 0.053 (-0.616; 0.722) | 0.876 | | |
| - Multi-component | -0.156 (-0.711; 0.402) | 0.587 | | |
| **Supervision** | | | 0.486 | 0.0 |
| - Physio | ref | | | |
| - Trainer | 0.154 (-0.362; 0.669) | 0.559 | | |
| **Frequency** | | | 0.521 | 0.0 |
| - 2 or 3 per wk | ref | | | |
| - 4 to 7 per wk | -0.165 (-0.669; 0.339) | 0.521 | | |
| **Duration of intervention** | | | 0.956 | 0.0 |
| - 0-12 wk | ref | | | |
| - 13-24 wk | 0.103 (-0.572; 0.778) | 0.765 | | |
| - > 24 wk | 0.008 (-0.404; 0.420) | 0.971 | | |
| **Intensity** | | | 0.957 | 0.0 |
| - Low | ref | | | |
| - Moderate | -0.025 (-0.489; 0.439) | 0.915 | | |
| - Vigorous | 0.044 (-0.470; 0.558) | 0.866 | | |

CI: Confidence Intervals; Ref = Reference subgroup for comparison; $R^2$ = amount of heterogeneity accounted for

### Figure S1: Overall meta-analysis (Follow-up)

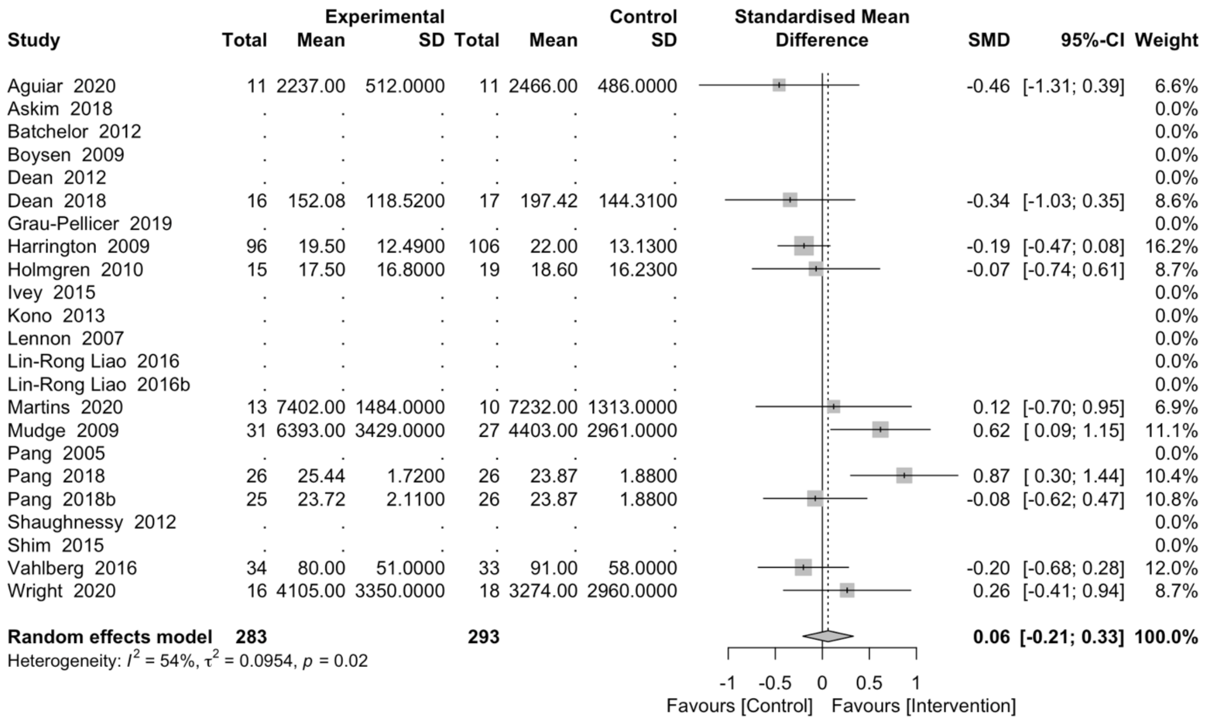


### Figure S2: Risk of bias

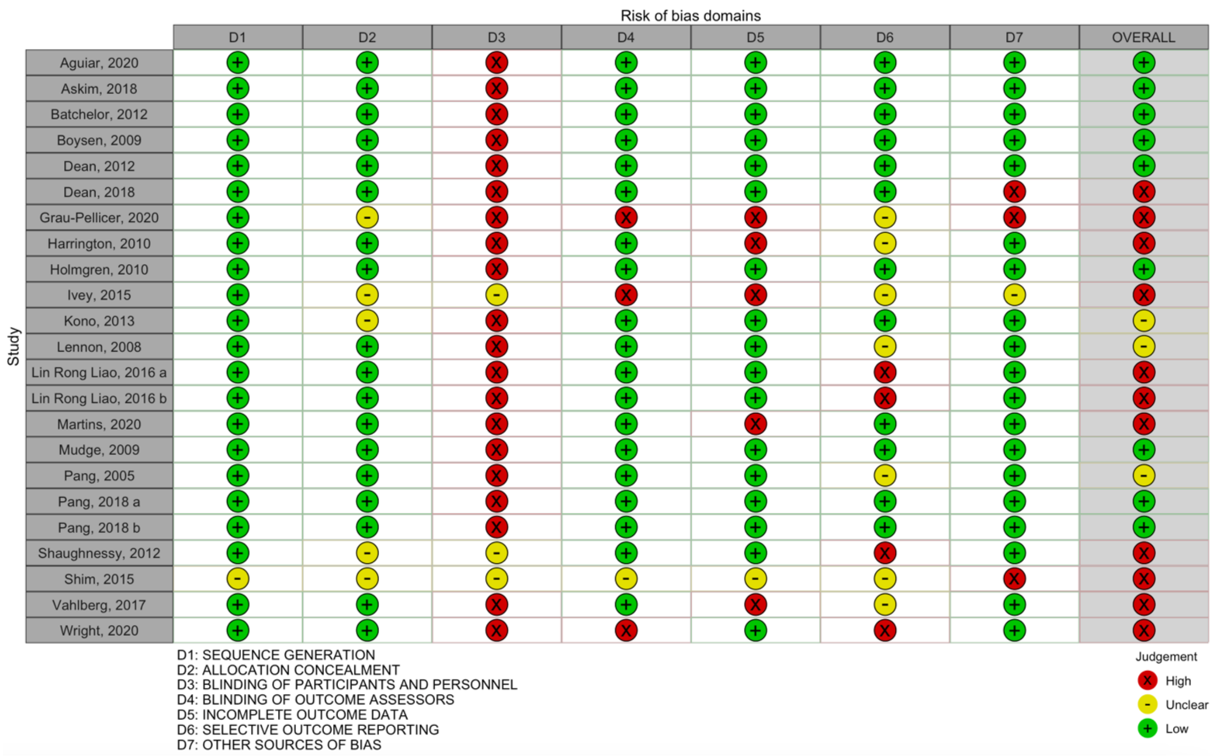

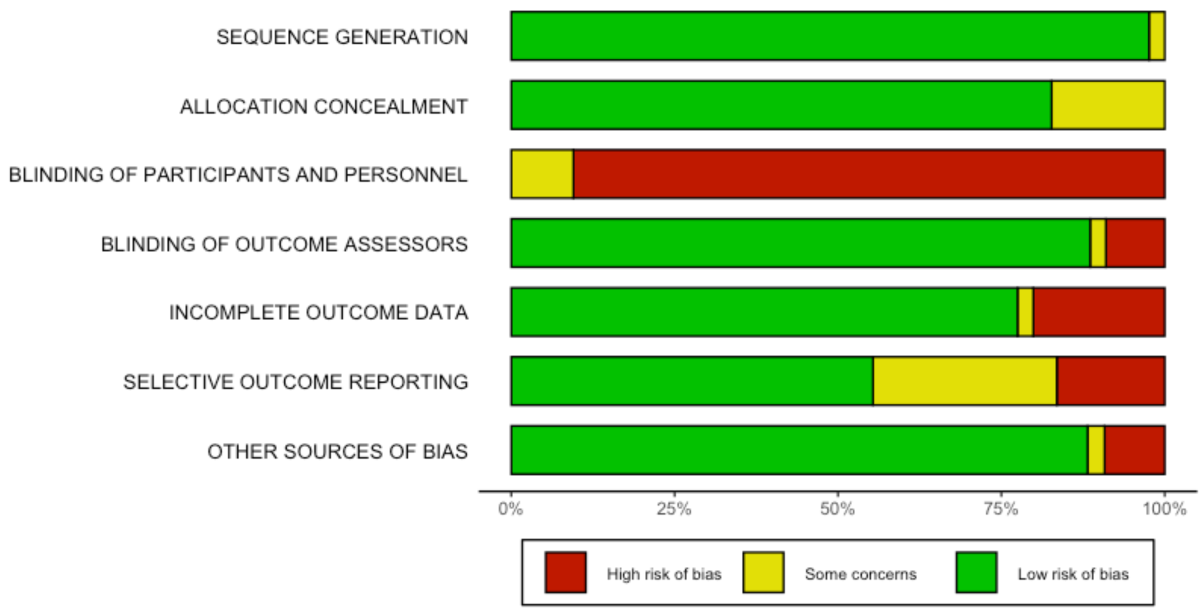
SEQUENCE GENERATION
ALLOCATION CONCEALMENT
BLINDING OF PARTICIPANTS AND PERSONNEL
BLINDING OF OUTCOME ASSESSORS
INCOMPLETE OUTCOME DATA
SELECTIVE OUTCOME REPORTING
OTHER SOURCES OF BIAS
0%
25%
50%
75%
100%
High risk of bias
Some concerns
Low risk of bias
